# Automated Dentate Nucleus Segmentation from QSM Images Using Deep Learning

**DOI:** 10.1101/2024.06.17.24308662

**Authors:** Diogo H Shiraishi, Susmita Saha, Isaac M Adanyeguh, Sirio Cocozza, Louise A Corben, Andreas Deistung, Martin B Delatycki, Imis Dogan, William Gaetz, Nellie Georgiou-Karistianis, Simon Graf, Marina Grisoli, Pierre-Gilles Henry, Gustavo M Jarola, James M Joers, Christian Langkammer, Christophe Lenglet, Jiakun Li, Camila C Lobo, Eric F Lock, David R Lynch, Thomas H Mareci, Alberto R M Martinez, Serena Monti, Anna Nigri, Massimo Pandolfo, Kathrin Reetz, Timothy P Roberts, Sandro Romanzetti, David A Rudko, Alessandra Scaravilli, Jörg B Schulz, S H Subramony, Dagmar Timmann, Marcondes C França, Ian H Harding, Thiago J R Rezende, TRACK-FA Neuroimaging Consortium

**Author notes:** **Corresponding author:** Dr. Thiago J R Rezende Department of Neurology, School of Medical Sciences, University of Campinas (Unicamp) Neuroimaging Laboratory Rua Vital Brasil, 89-99 Cidade Universitária “Zeferino Vaz” Campinas, São Paulo, Brazil Zip code: 13083-888. Contributed equally (Diogo H Shiraishi, Susmita Saha). Senior principal investigator (Ian H Harding, Thiago J R Rezende). see Acknowledgments for collaborators (TRACK-FA Neuroimaging Consortium).

## Abstract

**Purpose:** To develop a dentate nucleus (DN) segmentation tool using deep learning (DL) applied to brain quantitative susceptibility mapping (QSM) images.

**Materials and Methods:** Brain QSM images from 132 healthy controls and 170 individuals with cerebellar ataxia or multiple sclerosis were collected from nine different datasets worldwide for this retrospective study. Manual delineation of the DN (gray matter and white matter hilus) was first undertaken by experienced raters with a robust quality control process. Performance of automated segmentation was compared following training using several DL architectures. A two-step approach was implemented, composed of a localization model followed by DN segmentation.

**Results:** The manual tracing protocol produced ground-truth data with high intra-rater (average ICC 0.906) and inter-rater reliability (average ICC 0.776). Initial DL architecture exploration indicated that the nnU-Net framework performed best. The two-step localization plus segmentation pipeline achieved a Dice score of 0.898±0.031 and 0.894±0.036 for left and right DN, respectively. In external validation, our algorithm outperformed the leading existing automated tool (left/right DN Dice 0.863±0.038/0.843±0.066 vs. 0.568±0.222/0.582±0.239). The model demonstrated generalizability across unseen datasets during the training step. The measures showed a superior correlation index with manual annotations and performed well in both isotropic and anisotropic QSM datasets.

**Conclusion:** We provide a model that accurately and efficiently segments the DN from brain QSM images. The model can be readily deployed for use in observational, natural history, and treatment trials for biomarker discovery.

## 2. Introduction

The dentate nuclei (DN) are the largest of the deep cerebellar nuclei and are the primary efferent stations of the human cerebellum. The DN are primarily innervated by Purkinje cells of the lateral hemispheres of the cerebellar cortex and give rise to the major ascending cerebellar output pathways, including the dentato-rubral and dentato-thalamo-cortical tracts^1^. The DN are divided into motor and non-motor functional subregions with distinct profiles of disynaptic axonal innervation (*via* the thalamus) to cerebral cortices^1^. These cerebral regions, in turn, innervate corresponding subregions of the cerebellar cortex *via* descending disynaptic projections that pass through the pontine nuclei, forming the cerebro-cerebellar circuitry^2^.

Cerebro-cerebellar systems, with the DN as a key hub, are implicated in a broad array of motor, cognitive, social, and language functions^3^. Abnormalities in the DN may therefore contribute to disruption in large-scale brain networks involved in a broad array of behavioral deficits. Abnormal functional connectivity between the DN and brain cortical areas has been reported in patients with Alzheimer’s disease, autism, and schizophrenia^4–6^. Furthermore, neuropathological studies have provided evidence that the DN plays a central role in the pathogenesis of several cerebellar diseases, especially in the inherited cerebellar ataxias^7^.

Despite the established importance of the DN in cerebro-cerebellar loops, and growing evidence of involvement in brain diseases, direct *in vivo* quantitative investigations of the structure of the DN and other deep cerebellar nuclei using neuroimaging in humans are scarce^8–11^. Such investigations have been particularly challenging due to the tissue properties of these nuclei that make them invisible or poorly defined using standard MRI sequences, such as T1-weighted and T2-weighted images. Susceptibility-weighted MRI (SWI) offers a limited solution, allowing for visualization of the DN due to their high iron content^12–14^. However, SWI is a qualitative technique that is primarily used for the clinical detection of vascular abnormalities and microangiopathies^15^. Although useful, SWI has several limitations, including its non-quantitative nature and distortion of tissue boundaries due to blooming effects and non-local phase contributions of the iron deposits on the tissues. These limitations have largely been overcome through the development of quantitative susceptibility mapping (QSM)^16^. QSM allows for more precise mapping of the anatomy and a more direct link to the underlying iron concentration^17^, providing opportunities for direct, quantitative evaluation of DN structure and microstructure in clinical populations^8,11^.

QSM has been employed to demonstrate robust and/or progressive changes in the DN in people with inherited cerebellar ataxias, including Friedreich’s ataxia (FRDA)^11,18^ and spinocerebellar ataxias (SCA)^8^. Changes in the structure and susceptibility of DN related to healthy aging and other movement disorders, including Parkinson’s disease and essential tremor, have also recently been examined^9,19,20^. Taken together, these studies demonstrate the utility of QSM in the neuroimaging toolkit for examining the DN in health and disease, and motivate investigation of DN changes in other neurological, developmental, and psychiatric diseases that impact cerebellar circuitry.

Although QSM holds great promise for quantifying and tracking DN changes in disease, a major roadblock to-date in undertaking large-scale and reliable QSM investigations of the DN in patient cohorts has been the reliance on manual, hand-drawn segmentations. In order to overcome this limitation, fully automated tools are necessary. The MRICloud toolkit^21,22^ provided the first (and currently, to our knowledge, only) publicly available automated DN segmentation tool. However, MRICloud has not been trained on data with DN pathology. In this work, we address these limitations by utilizing data from healthy subjects and cerebellar ataxia or multiple sclerosis (MS) patient cohorts acquired at multiple imaging centers using different acquisition protocols to develop an optimized and generalizable deep-learning analytical tool for segmenting the DN using QSM images. This tool can be readily deployed in observational, natural history, and treatment trials.

## 3. Materials and Methods

### 3.1. Data

Multi-echo gradient-recalled echo MRI data was acquired using four different MRI protocols implemented across ten imaging centers around the world using 3 Tesla Philips or Siemens MRI scanners (Tables 1 and 2). The collected research data was de-identified at each source, ensuring adherence to a data pipeline free from personal health information.

**Table 1.** Acquisition protocols for each dataset.

| Dataset | Scanner | Sequence | TR (ms) | TE1 (ms) | ΔTE (ms) | # of echoes | FoV (mm) | Image matrix (voxels) | Voxel size (mm) | Acquisition time |
| --- | --- | --- | --- | --- | --- | --- | --- | --- | --- | --- |
| Aachen<br>Campinas<br>CHoP<br>Melbourne<br>Florida<br>Minnesota | 3T Siemens Skyra<br>3T Siemens Prisma<br>3T Philips Ingenia | GRE | 27 | 3.7 | 6 | 4 | 220 x 220 x 176 | 208 x 256 x 176 | 0.86 iso | 7' 22" |
| Naples | 3T Siemens Magnetom Trio | GRE | 32 | 7.38 | 14.76 | 2 | 230 x 194 x 160 | 378 x 448 x 160 | 0.51 x 0.51 x 1.0 | 9' 53" |
| IMAGE-FRDA | 3T Siemens Skyra | GRE | 30 | 7.38 | 14.76 | 2 | 230 x 230<br>160 axial slices | 232 x 256 x 160 | 0.90 iso | 11' 30" |
| INFLAM-FRDA | 3T Siemens Biograph | GRE | 31 | 5.70 | 5.27 | 5 | 230 x 230<br>104 axial slices | 384 x 384 x 104 | 0.60 x 0.60 x 1.2 | 6' 50" |

GRE: gradient recalled echo; iso: isotropic; TR: repetition time; TE: echo time; TE1: first echo time; FoV: field of view; iso: isotropic.

**Table 2.** Subject demographics for each dataset.

|  | Aachen, Campinas,<br>CHoP, Melbourne,<br>Florida, Minnesota |  | Naples |  |  | IMAGE-FRDA |  | INFLAM-FRDA |  |
| --- | --- | --- | --- | --- | --- | --- | --- | --- | --- |
|  | Controls | FRDA | Controls | FRDA | MS | Controls | FRDA | Controls | FRDA |
| **Subjects** | 44 | 98 | 48 | 12 | 16 | 31 | 30 | 9 | 14 |
| **Children (<18 years)** | 14 | 31 | 0 | 3 | 0 | 0 | 0 | 0 | 0 |
| **Age (mean±SD)** | 23.6±7.7 | 23.9±8.7 | 37.8±12.7 | 32.6±15.8 | 46.8±5.6 | 37.6±13.1 | 35.7±12.2 | 28.6±5.8 | 27.8±7.6 |
| **Sex (M/F)** | 24/20 | 49/49 | 19/29 | 4/8 | 5/11 | 16/15 | 17/13 | 8/1 | 12/2 |

FRDA: Friedreich's ataxia; MS: multiple sclerosis.

The dataset included a total of 132 healthy controls, 154 individuals with FRDA, and 15 people with MS, as described in Table 2. Follow-up scans at 12 months were available from 55 of these participants. Images of FRDA and MS patients were selected to account for anatomical variability in DN caused by neurodegeneration throughout the course of the diseases. The corresponding MRI acquisition protocols have been previously published^11,23,24^ and are summarized in Table 1.

A sample QSM image for each dataset is presented in Figure 1, and Figure 2 provides an overview of the data workflow. The QSM images were reconstructed using the JHU/KKI QSM^21,25^ and STI Suite (https://people.eecs.berkeley.edu/~chunlei.liu/software.html) toolboxes, using Laplacian unwrapping to overcome phase aliasing, V-SHARP for background field removal^26^ and either MEDI^27,28^ or iLSQR^29^ for field-to-susceptibility reconstruction.

**Figure 1.**
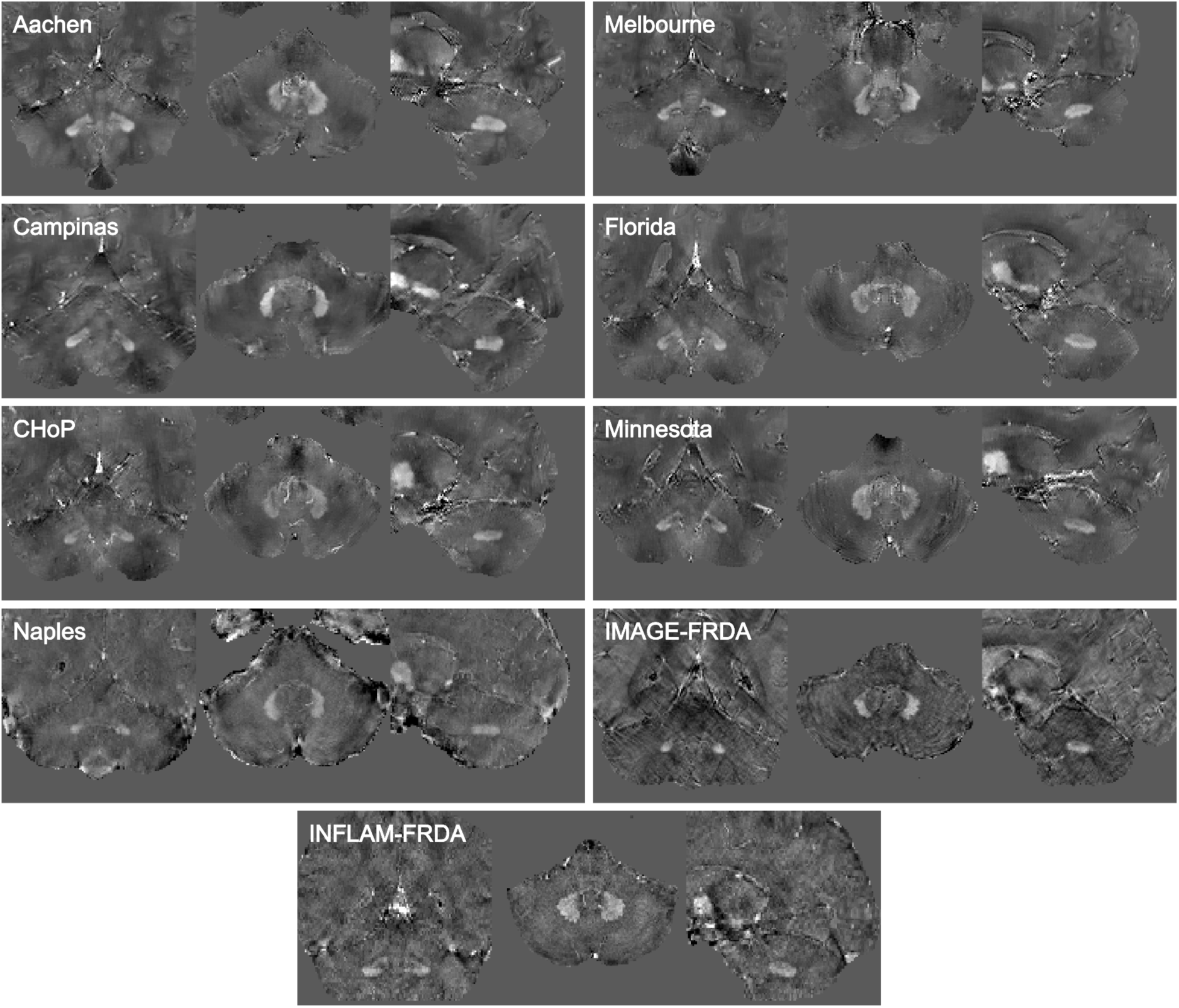
Image samples for each dataset. Randomly selected. Images were cropped focusing on the cerebellum, voxel intensities were normalized using z-score to improve visualization.

**Figure 2.**
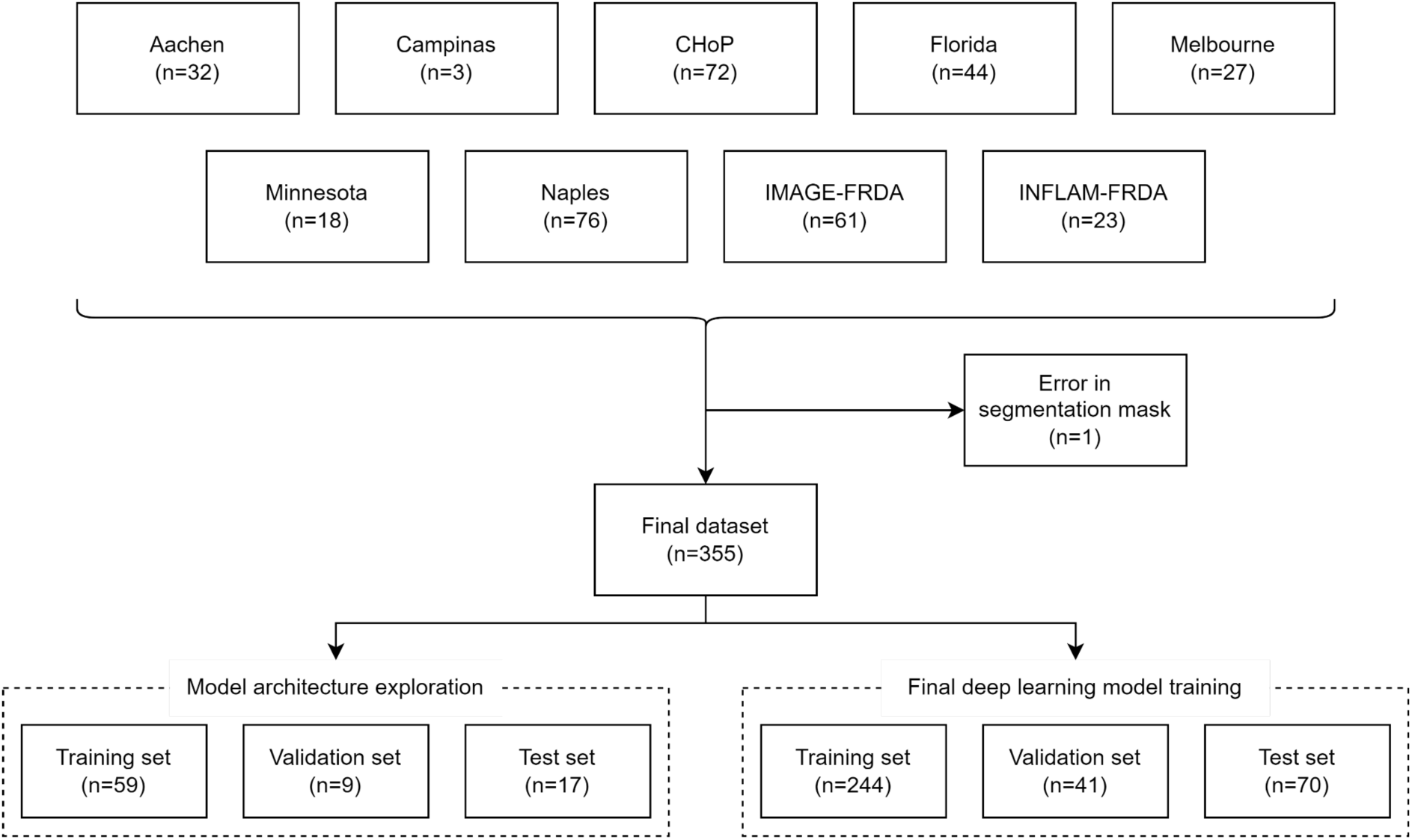
Diagram details the inclusion of image datasets into the study.

To train the deep learning (DL) models, we divided the available data into 70/10/20 % proportions for training, validation, and test sets, respectively. We applied stratified sampling, creating groups that maintain the relative distributions by MRI acquisition center, while also keeping images belonging to an individual in just one set, minimizing the possibility of data leakage.

The ethics committee or institutional review board (IRB) respective to each project data source or site approved the use or ethics waiver for this retrospective study, and all participants provided written informed consent prior to original data collection. The TRACK-FA steering committee approved the data use, and IRB reference numbers were previously published^23^ (Monash Health Human Research Ethics Committee: RES-20-0000-139A; Children’s Hospital of Philadelphia: IRB 20-017611; University of Minnesota: IRB STUDY00009047; University of Florida: IRB202000399; RWTH Aachen University: EK195/20; University of Campinas (CAAE NO): 83241318.3.1001.5404; McGill IRB Approved Project Number: 2022-8676). Ethics approval was obtained independently for the remaining studies, respectively: Ethical Committee “Carlo Romano” of the University of Naples “Federico II” (Naples A: 209/13, Naples B: 47/15), Monash University Human Research Ethics Committee (IMAGE-FRDA: 13201B, INFLAM-FRDA: 7810), and University of Minnesota IRB (1210M22281).

### 3.2. Ground Truth Segmentation

The DN were manually traced on each QSM image to establish the ground truth dataset using MRIcron (https://www.nitrc.org/projects/mricron), ITK-SNAP^30^, or FSLeyes^31^ annotation software. The contrast threshold was set at a constant range (−0.1 to 0.2 ppm) for all images to minimize bias in edge detection within and across raters.

The segmentations included the full 3D region of hyperintensity, including both the outer gray matter ribbon and the central white matter hilum of the DN (Figure 3). Manual tracing was performed in all three planes (axial, coronal, and sagittal) to ensure a smooth surface was generated and neighboring nuclei (e.g., emboliform) were not included. The left DN (LDN) and right DN (RDN) were uniquely segmented on each image (Figure 3).

**Figure 3.**
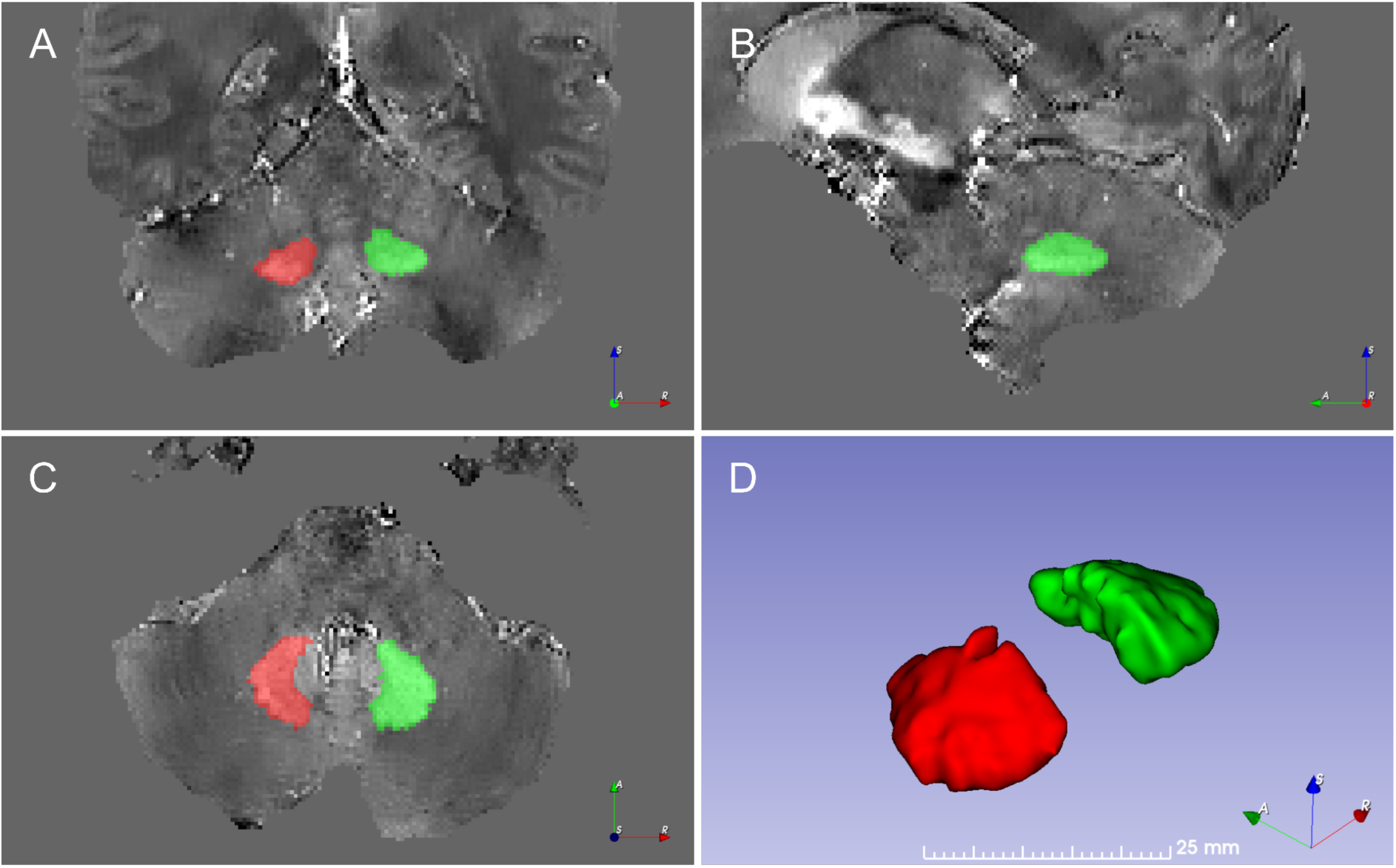
Manually segmented mask delineating the left dentate nucleus (LDN) in red and the right dentate nucleus (RDN) in green from a sample QSM dataset, healthy participant. Coronal, sagittal, and axial views as well as shaded-surface display are shown from A - D, respectively.

Each image was manually segmented by one of three experienced raters (SS, SC, or IHH), blinded to the disease pathology. Inter-rater and intra-rater reliability was assessed using a sample of 9 randomly selected images, stratified by acquisition center. These images were duplicated and randomly shuffled so that the rater was unaware of the order or the repetition. All three raters segmented this set of 18 images. Intra-rater and inter-rater variability were calculated using Dice score, Hausdorff Distance (HD), and intraclass correlation coefficient (ICC) metrics.

### 3.3. Deep Learning Architectures

A number of artificial intelligence approaches for image segmentation are available. In the DL domain, convolutional neural network (CNN) architectures are typically used to process image data. For image segmentation tasks, a great number of studies rely on U-Nets^32^ and their 3D variation, 3D U-Net^33^, both based on CNNs. Numerous variations of these architectures have been reported in the literature. The no-new-Net (nnU-Net) is considered one of the state-of-the-art medical imaging segmentation frameworks, capable of dealing with image sets comprising different domains^34^. The nnU-Net does not implement a rigorous complex architecture, instead relying on a conventional U-Net with deep supervision^35^. More recently, the scientific community introduced new architectures based on transformers^36^, which arose in the language sequence models domain. These include TransUNet^37^, UNETR^38^ and Swin UNETR^39^. In this work, we applied and contrasted the performance of three DL architectures, described below, to develop an optimal approach for automated DN parcellation.

### 3.4. 3D U-Net with Deep Supervision

The 3D U-Net^33^ is an extension of the original U-Net architecture^32^ to handle three-dimensional data. Designed for image segmentation tasks, the architecture is composed of a contracting path, which gradually downsamples the input image, and an expanding path, which upsamples the feature maps back to the original image size. Also, the network makes use of skip connections, allowing information to flow directly from the contracting path to the corresponding layer in the expanding path. This helps to preserve spatial information and improve the accuracy of the segmentation. Additionally, the 3D U-Net can leverage the context information in the whole 3D image, learning intricate and complex spatial relationships between the voxels in the image.

Deep supervision (DS)^35^ is a technique that adds extra output layers at different levels of the network. These output layers are trained to predict the final segmentation and guide the network in the earlier stages of training, providing additional guidance that helps improve segmentation accuracy. This allows gradients to be injected deeper into the network and facilitates the training of all layers in the network^34^. This architecture is also adopted by the nnU-Net framework.

### 3.5. Swin UNETR

The Shifted WINdows UNEt TRansformers (Swin UNETR) is a Swin transformer-based deep learning model designed for medical image segmentation^39^. Swin transformer^40^ implements a hierarchical vision transformer that computes self-attention locally in non-overlapping windows, enabling the model to capture long-range dependencies. In the hierarchical structure, patches are merged layer by layer, decreasing the dimension of feature maps at each stage. The cyclic-shifting implements the shifted window partitioning approach and introduces important cross-windows connections, overlapping with previous windows, while the number of patches remains fixed. These properties explain the linear computational complexity to image size, unlike to the quadratic in other transformed-based models, such as Vision Transformers^41^ (ViT), which would impose an issue of scalability for semantic segmentation, a dense prediction task. Swin transformers build hierarchical feature maps to obtain multi-resolution feature representations, making them suitable as backbone networks for various computer vision tasks. In Swin UNETR, the Swin transformer serves as the encoder and is integrated with a CNN-based decoder through skip connections in a U-shaped architecture.

### 3.6. nnU-Net

The nnU-Net is a deep learning framework for medical image segmentation that features automated configuration^34^. The core architecture of nnU-Net is a U-Net with DS, which is enhanced with a data-driven initialization strategy. This approach adapts the training to the specific characteristics of the input data, resulting in improved performance. The pipeline includes data augmentation and training across different configurations such as 2D U-Net, 3D U-Net at full resolution, and 3D U-Net cascade (a combination of a model trained on downsampled images followed by a refinement model at full resolution).

Additionally, the nnU-Net has the capability to apply post-processing steps, such as “non-largest component suppression”, if it improves the results. It can also self-adjust hyperparameters to fit the GPU memory available during training.

This framework has been demonstrated to achieve state-of-the-art performance on various medical image segmentation tasks, leading to widespread adoption in the medical imaging community.

### 3.7. Augmentation Techniques

In order to improve model generalization capacity, we applied data augmentation techniques such as flipping, rotation, scaling, elastic deformation, and intensity scale and shift operations. These operations were randomly applied to each image during the training phase. Utilizing these transformations is an important strategy to artificially increase data variability, thereby preventing overfitting and boosting generalizability. To accomplish this, we selected MONAI^42^ augmentation implementations.

### 3.8. Deep Learning Segmentation Pipeline

The DL segmentation pipeline is presented in Figure 4. First, the QSM image is resampled to a common isotropic voxel spacing of 0.86 mm, consistent with the original resolution of the majority of the datasets.

**Figure 4.**
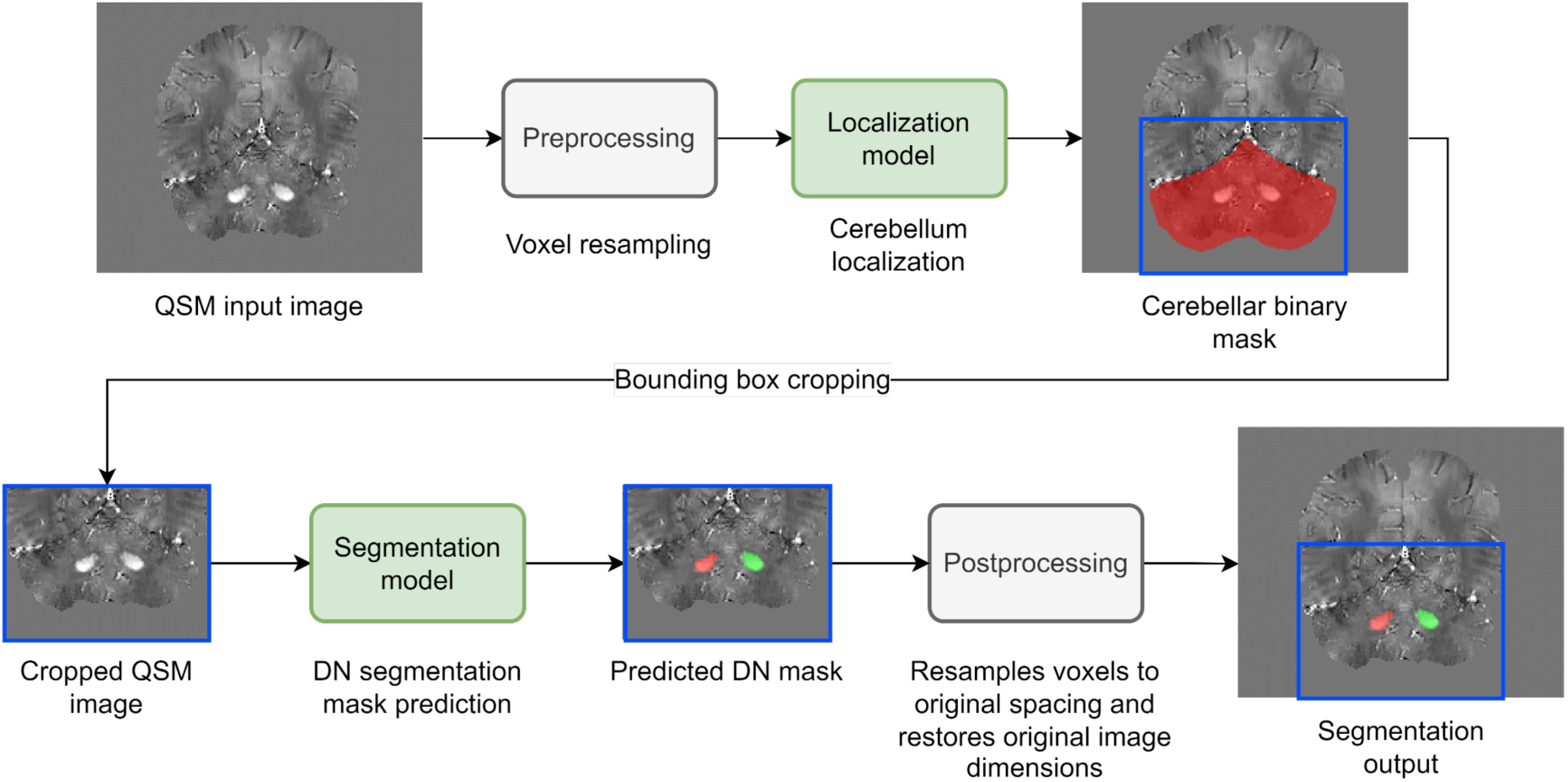
Deep learning QSM segmentation pipeline.

The second step in our pipeline is the application of the localization model, which is a deep learning network designed to identify the location of the cerebellum within a 3D MRI dataset. The localization model was trained with cerebellar masks obtained with ACAPULCO^43^ based on T1-weighted MRI of each subject. To ensure spatial alignment of the cerebellum masks with the QSM images, the T1w data were registered to the corresponding QSM data using ANTs toolkit^44^, and the resulting transformation matrix was applied to the cerebellum mask. The localization model will therefore output a cerebellar mask. The centroid of the mask is determined and used to place a bounding box cropping around the cerebellum, ensuring the preservation of the entire structure without any loss or truncation, thereby constraining and reducing the spatial extent of the region of interest (RoI). This approach allows the retention of contextual anatomical references.

The third step was the segmentation stage. The output of this model was the binary mask for each dentate nucleus label (left and right). Subsequently, another resampling process is performed to provide the predicted DN segmentation mask in the original voxel spacing and in the same dimensions as the input image.

### 3.9. Statistical Analysis

The Sørensen-Dice similarity coefficient (DSC), also known as the Dice score coefficient^45,46^, is a widely used metric for assessing segmentation tasks. It is noteworthy that the Dice score, which measures the overlap between two binary segmentation masks, considers both size and localization agreement^47^ that is, both volumetric and anatomic characteristics. The Dice score is defined as follows:

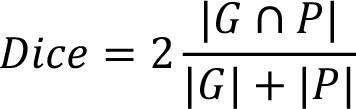

where |𝐴| is the cardinality of set 𝐴. 𝐺 and 𝑃 represent the ground truth segmentation mask and the model prediction segmentation, respectively. This metric ranges from 0 to 1, with a value of 1 indicating a perfect agreement. The Dice score was our main performance metric, and it was also part of the loss metric for model training in the form of Dice loss:

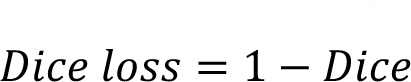

Additionally, other segmentation metrics were evaluated. The Hausdorff Distance^48^ and Average Hausdorff Distance (AHD)^49^ are relevant for boundary quality assessment.

AHD calculates the average distance between points in both masks instead of considering only the maximum distance as in HD. Also, Intersection over Union (IoU) or Jaccard index^50^, which is similar to Dice, was evaluated. Finally, volume similarity^49^ was assessed to measure the quantitative volume agreement, ignoring shape, position, and quality information. To compare metrics between models, we used the Wilcoxon signed-rank test. Kolmogorov-Smirnov test was employed to assess normality in continuous variables. Pearson correlation coefficients were chosen to evaluate relationships within variables. We measured the reliability of measurements from multiple raters using the intraclass correlation coefficient (ICC) and evaluated the results (ICC<0.5 poor, 0.5≤ICC<0.75 moderate, 0.75≤ICC<0.9 good, and ICC≥0.9 excellent reliability)^51^. ICC confidence intervals were calculated to provide a range within which the ICC is likely to fall, typically with 95% confidence. Statistical analysis was conducted using Python 3.10.8, R 4.1.1, and SPSS 27.0.1.0. P-values < 0.05 were considered statistically significant.

### 3.10. Comparison with other available methods

In order to compare our results with the leading available automated DN segmentation solution, we processed all our test images through the MRICloud web-based service^21,22^ using its susceptibility multi-atlas tool for segmentation of QSM images.

The MRICloud pipeline requires both coregistered QSM and skull-stripped T1 images in ANALYZE file format. We used ANTs (SyNQuick) for co-registration, FSL^52^ Brain Extraction Tool (BET) for skull-stripping, and FSL for file conversion between NIfTI and ANALYZE formats. MRICloud jobs for the segmentation model architecture comparison section were executed between January 5 and January 7, 2023, and then again from March 9 to March 31, 2024, for the external validation datasets.

### 3.11. External Validation

In order to assess the performance of our DN segmentation model on unseen data, 21 external datasets of 10 participants were acquired from three new imaging sites: n=4 healthy participants from Instituto Neurologico “Carlo Besta”, Italy; n=2 MS patients from the Medical University of Graz, Austria; and n=3 healthy participants and n=1 FRDA patient from McGill University. QSM maps were derived for all participants using both the JHU/KKI QSM v3.0^21,53^ and SEPIA v1.2.2.6^54^ toolboxes, resulting in two images per participant for Carlo Besta and McGill datasets. Three reacquired Carlo Besta anisotropic QSM images processed with SEPIA were added. An experienced rater (SS) manually segmented the right and left DN, and the data was processed through both our models and MRICloud to compare their performances. For SEPIA processing, 3D best path^55^ was used for phase unwrapping and MEDI non-linear fit for echo phase combination, Laplacian Boundary Value (LBV)^56^ approach for background magnetic field removal, and quantitative susceptibility maps were obtained using STreaking Artifact Reduction for QSM (Star-QSM)^57^.

The institutional ethics committee respective to each project approved their study (Fondazione IRCCS Istituto Neurologico “Carlo Besta”: 42/2017 07/06/2017; Medical University of Graz local ethics-committee: 31-432ex18/191264-2019).

## 4. Results

### 4.1. Intra-Rater and Inter-Rater Reproducibility Results

All raters demonstrated good to excellent intra-rater reliability (ICC > 0.8; Table 3), and moderate (ICC > 0.5) to good inter-rater agreement (ICC > 0.75) when evaluated in pairs^51^ (Table 4). Analyzing all three raters at once, the ICC is 0.763, 95% CI [0.541, 0.897] for LDN and 0.675, 95% CI [0.173, 0.883] for RDN. Dice and HD additionally support strong reliability, evaluating segmentation overlapping and annotation deviations. Volume similarity metric, comparing the segmented volumes (mm^3^), indicates that absolute differences within and between raters are marginal. The results are shown in Tables 3 and 4.

**Table 3.** Intra-rater reproducibility results (mean±SD).

| Metric | Rater 1 |  | Rater 2 |  | Rater 3 |  |
| --- | --- | --- | --- | --- | --- | --- |
|  | LDN | RDN | LDN | RDN | LDN | RDN |
| **Dice** | 0.884±0.025 | 0.888±0.027 | 0.904±0.023 | 0.918±0.024 | 0.892±0.014 | 0.912±0.010 |
| **HD** | 3.276±1.174 | 3.491±1.169 | 2.397±0.837 | 2.109±0.879 | 2.324±0.394 | 1.745±0.256 |
| **Volume similarity** | 0.943±0.037 | 0.961±0.029 | 0.964±0.023 | 0.963±0.029 | 0.969±0.016 | 0.980±0.015 |
| **ICC** | 0.839 | 0.849 | 0.939 | 0.902 | 0.930 | 0.977 |
| **[95% CI]** | [0.258, 0.965] | [0.440, 0.964] | [0.306, 0.989] | [0.640, 0.977] | [0.735, 0.984] | [0.905, 0.995] |

LDN: left dentate nucleus, RDN: right dentate nucleus, HD: Hausdorff distance, ICC: intraclass correlation coefficient, CI: confidence interval.

**Table 4.** Inter-rater reproducibility results (mean±SD).

| Metric | Raters |  |  |  |  |  |
| --- | --- | --- | --- | --- | --- | --- |
|  | 1 and 2 |  | 2 and 3 |  | 1 and 3 |  |
|  | LDN | RDN | LDN | RDN | LDN | RDN |
| **Dice** | 0.852±0.040 | 0.864±0.027 | 0.852±0.032 | 0.855±0.025 | 0.872±0.020 | 0.870±0.028 |
| **HD** | 3.361±1.325 | 3.569±1.021 | 3.416±1.154 | 3.573±1.230 | 2.946±1.266 | 3.867±1.451 |
| **Volume similarity** | 0.931±0.055 | 0.944±0.041 | 0.918±0.040 | 0.897±0.041 | 0.950±0.026 | 0.933±0.039 |
| **ICC** | 0.751 | 0.740 | 0.709 | 0.599 | 0.845 | 0.731 |
| **[95% CI]** | [0.451, 0.899] | [0.298, 0.905] | [0.268, 0.890] | [-0.079, 0.886] | [0.376, 0.951] | [-0.048, 0.925] |

LDN: left dentate nucleus, RDN: right dentate nucleus, HD: Hausdorff distance, ICC: intraclass correlation coefficient, CI: confidence interval.

### 4.2. Localization Model

The model for the cerebellum localization features a 3D U-Net model with feature channels ranging from 16 to 256, doubling at each subsequent level. Levels combine two 3×3×3 convolutions, followed by instance normalization^58^ and Parametric Rectified Linear Unit (PReLU)^59^ activations. An AdamW optimizer^60^ with learning rate = 3×10^-4^, weight decay = 1×10^-5^, β1 = 0.9, β2 = 0.999, and ɛ = 1×10^−8^. A learning rate scheduler reduced the learning rate by a factor of 0.5 if no improvement in loss was observed for 20 epochs. The batch size was configured to 1, and training was limited to 400 epochs. After 105 epochs, the model training was interrupted by the early stopping scheduler, after 30 epochs without a decrease in Dice loss. Consequently, the model checkpoint at epoch 75, which performed best on the validation set, was saved. The trained model showed a high Dice score, 0.918±0.030, when compared to the ground truth. After running inference for all available QSM images, they were cropped to 128×96×96, considering the centroid of the mask as the crop center for the enclosing region of interest. The localization model inference runs in less than five seconds when executed on CPU-only hardware and provides the model with as much spatial context as possible, while taking into account computational resources constraints.

### 4.3. Segmentation Model Architectures Comparison

All the results presented below refer to metrics calculated on the test set (17 images) from the model experimentation set (Figure 2). The performance of three model architectures (3D U-Net with DS, Swin UNETR architectures, and nnU-Net framework) were compared with MRICloud (Table 5). The nnU-Net architecture provided the best performance across most comparison metrics (Table 5), and all DL architectures were substantially superior to MRICloud (Dice score comparison, p<0.0001 in all cases; Figure 5). Jaccard index and AHD also indicate an advantage for nnU-Net, considering overlap and mask similarity. While the Hausdorff distance is lower in the U-Net with DS model, the average values for the two dentate nuclei are close to those of nnU-Net. Finally, despite U-Net with DS having the best average volume similarity, the nnU-Net standard deviation is lower, which is preferable.

**Table 5.** Trained models and MRICloud performance metrics (mean±SD) for left DN (LDN) and right DN (RDN) on the experimentation set.

| Model | Dice |  | Jaccard |  | HD |  | AHD |  | Volume similarity |  |
| --- | --- | --- | --- | --- | --- | --- | --- | --- | --- | --- |
|  | LDN | RDN | LDN | RDN | LDN | RDN | LDN | RDN | LDN | RDN |
| U-Net with DS | 0.889±0.021 | 0.895±0.023 | 0.802±0.033 | 0.810±0.037 | **2.514±0.749** | 2.361±0.654 | 0.117±0.024 | 0.112±0.027 | **0.965±0.021** | **0.963±0.026** |
| Swin UNETR | 0.890±0.022 | 0.896±0.021 | 0.803±0.035 | 0.812±0.034 | 2.887±0.885 | 2.495±0.550 | 0.120±0.032 | 0.111±0.024 | 0.962±0.023 | 0.953±0.029 |
| nnU-Net | **0.896±0.020** | **0.900±0.021** | **0.811±0.032** | **0.819±0.034** | 2.518±0.915 | **2.198±0.534** | **0.112±0.025** | **0.106±0.025** | 0.964±0.018 | 0.958±0.023 |
| MRICloud | 0.725±0.074 | 0.760±0.053 | 0.573±0.090 | 0.616±0.068 | 6.302±2.611 | 5.736±1.725 | 0.445±0.187 | 0.367±0.125 | 0.850±0.086 | 0.877±0.074 |

DN: dentate nucleus; LDN: left DN; RDN: right RN; DS: deep supervision; HD: Hausdorff distance; AHD: average Hausdorff distance.

**Figure 5.**
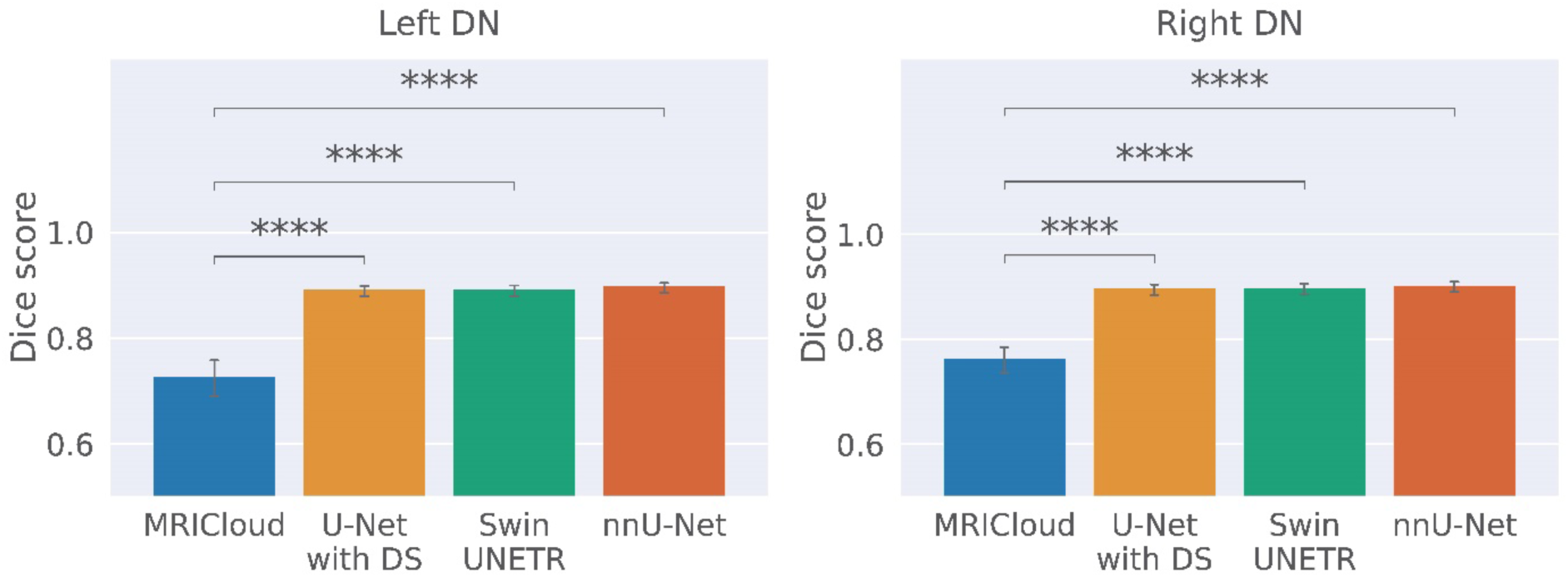
Bar plots of the trained segmentation models and MRICloud. All metrics of the segmentation model variants are statistically significantly higher than the MRICloud results. ns: non-significant; * p < 0.05; ** p < 0.01; *** p < 0.001; **** p < 0.0001. DN: dentate nucleus, DS: deep supervision.

After the extensive exploration of model architectures, the nnU-Net framework was selected for our study.

### 4.4. Final Segmentation Model Outputs

The final trained model (nnU-Net) with the entire dataset resulted in metrics on the test set presented in Table 6, including those for the control and patient groups individually. Samples of predicted segmentation masks for each center are shown in Figure 6. In agreement with that, the predicted DN volumes highly correlate with the volumes of the ground truth annotations (Figure 7).

**Table 6.** Trained nnU-Net final model performance metrics (mean±SD) for left DN (LDN) and right DN (RDN) on the complete test set.

| Group | Dice |  | Jaccard |  | HD |  | AHD |  | Volume similarity |  |
| --- | --- | --- | --- | --- | --- | --- | --- | --- | --- | --- |
|  | LDN | RDN | LDN | RDN | LDN | RDN | LDN | RDN | LDN | RDN |
| All participants | 0.898±0.031 | 0.894±0.036 | 0.816±0.050 | 0.810±0.057 | 3.124±1.825 | 4.025±4.569 | 0.118±0.060 | 0.140±0.144 | 0.943±0.044 | 0.944±0.050 |
| Controls | 0.893±0.033 | 0.891±0.044 | 0.808±0.053 | 0.806±0.067 | 3.408±1.916 | 3.850±2.046 | 0.127±0.068 | 0.134±0.073 | 0.949±0.046 | 0.951±0.058 |
| Patients* | 0.901±0.029 | 0.896±0.032 | 0.821±0.048 | 0.813±0.051 | 2.932±1.783 | 4.144±5.747 | 0.112±0.055 | 0.144±0.179 | 0.940±0.043 | 0.939±0.044 |

\* FRDA and MS patients. FRDA: Friedreich's ataxia; MS: multiple sclerosis; DN: dentate nucleus; LDN: left DN; RDN: right RN; HD: Hausdorff distance; AHD: average Hausdorff distance.

**Figure 6.**
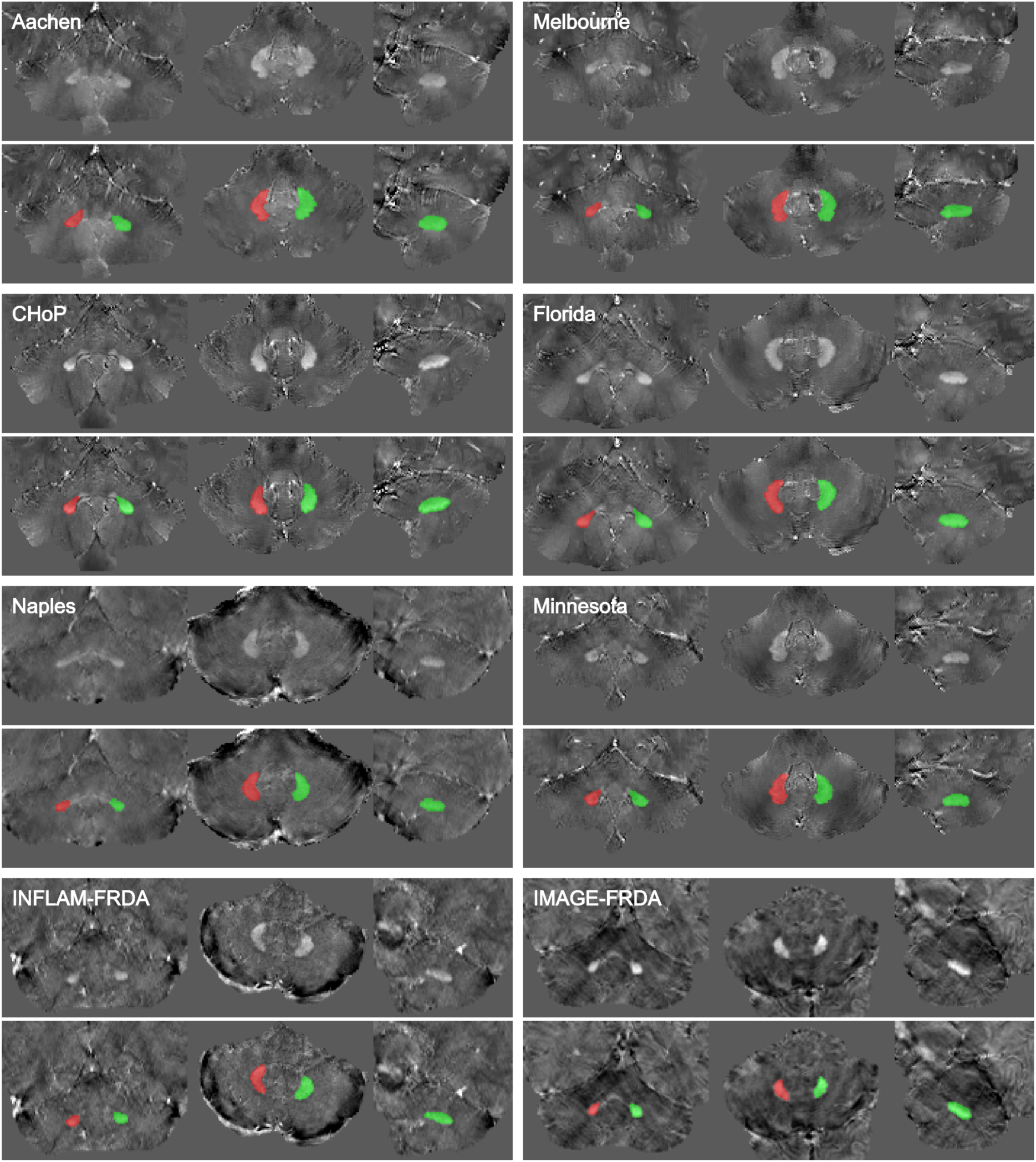
Overview of pipeline prediction examples for datasets of each center. QSM images without (top) and with the predicted segmentation mask as overlay (bottom) are presented in the coronal, axial, and sagittal planes, respectively. Left dentate nucleus is depicted in red and the right one in green.

**Figure 7.**
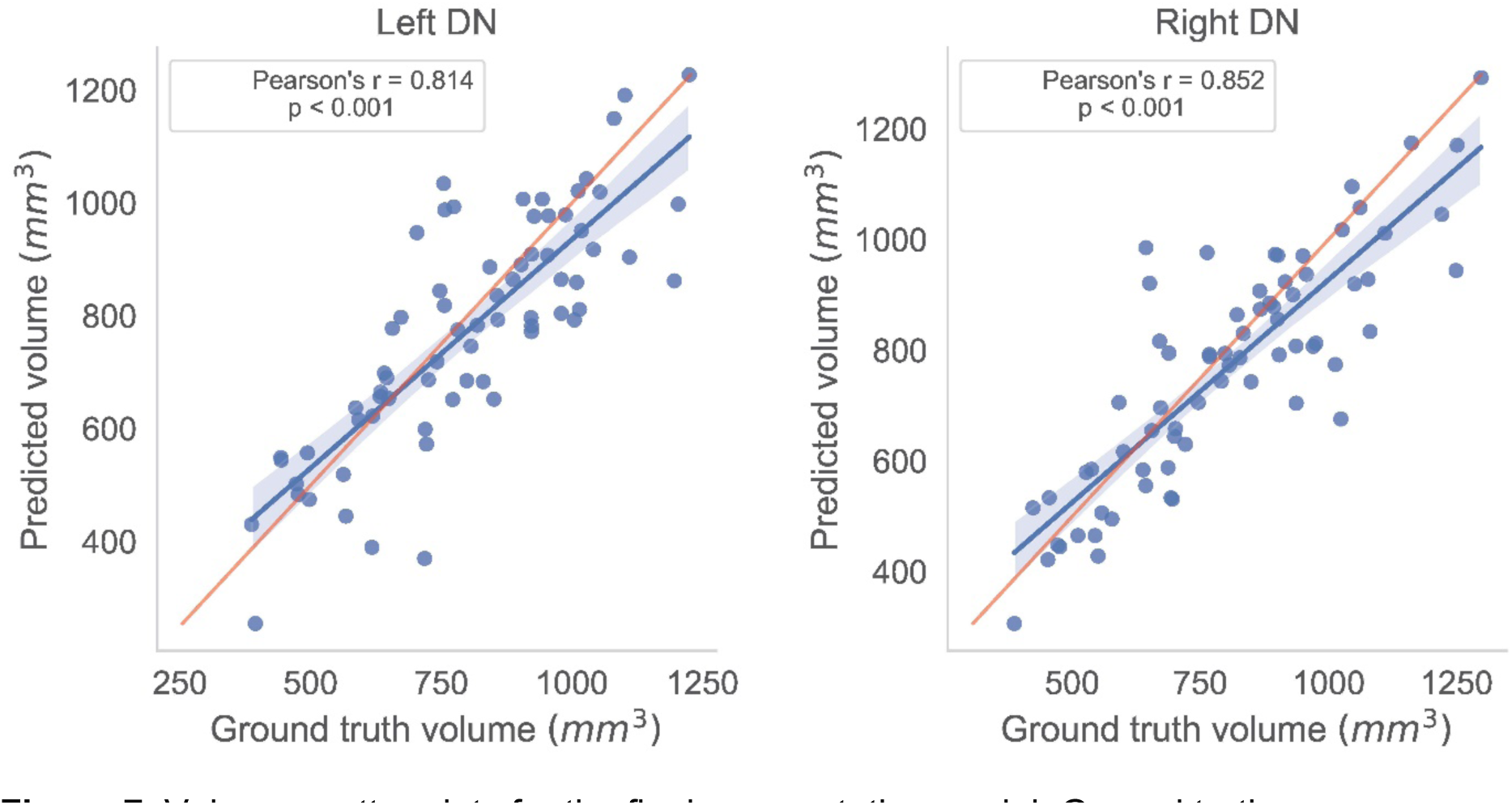
Volume scatter plots for the final segmentation model. Ground truth measures versus model predicted values. Each dot represents a prediction sample from the test set. Pearson correlation coefficients and p-values are indicated in each plot. The red line represents the line of identity, i.e., the expected perfect fit. DN: dentate nucleus.

### 4.5. External Validation

Our model performed satisfactorily on an external dataset of 21 images from three different sites, achieving Dice scores of 0.863±0.038 for LDN and 0.843±0.066 for RDN. In contrast, MRICloud demonstrated significantly lower performance for both LDN (Dice score: 0.568±0.222) and RDN (Dice score: 0.582±0.239). The observed performance difference in Dice metric was found to be statistically significant according to the Wilcoxon test for both LDN and RDN (p<0.05). Additionally, Pearson correlation between volumes predicted by the DL model (LDN: r=0.740, p<0.001; RDN: r=0.484, p=0.026) and those manually traced was significant for both DN sides (p<0.05), a result not observed with MRICloud outcomes (LDN: r=0.420, p=0.058; RDN: r=0.187, p=0.417). Interestingly, our model showed consistency across three external datasets when considering the individual QSM subsets per acquisition center. Furthermore, our segmentation pipeline demonstrated robust performance when evaluating susceptibility maps generated by the alternative QSM reconstruction method (Star-QSM, see section 3.11), yielding high Dice scores both overall and individually for each site (Tables 7 and 8).

**Table 7.**
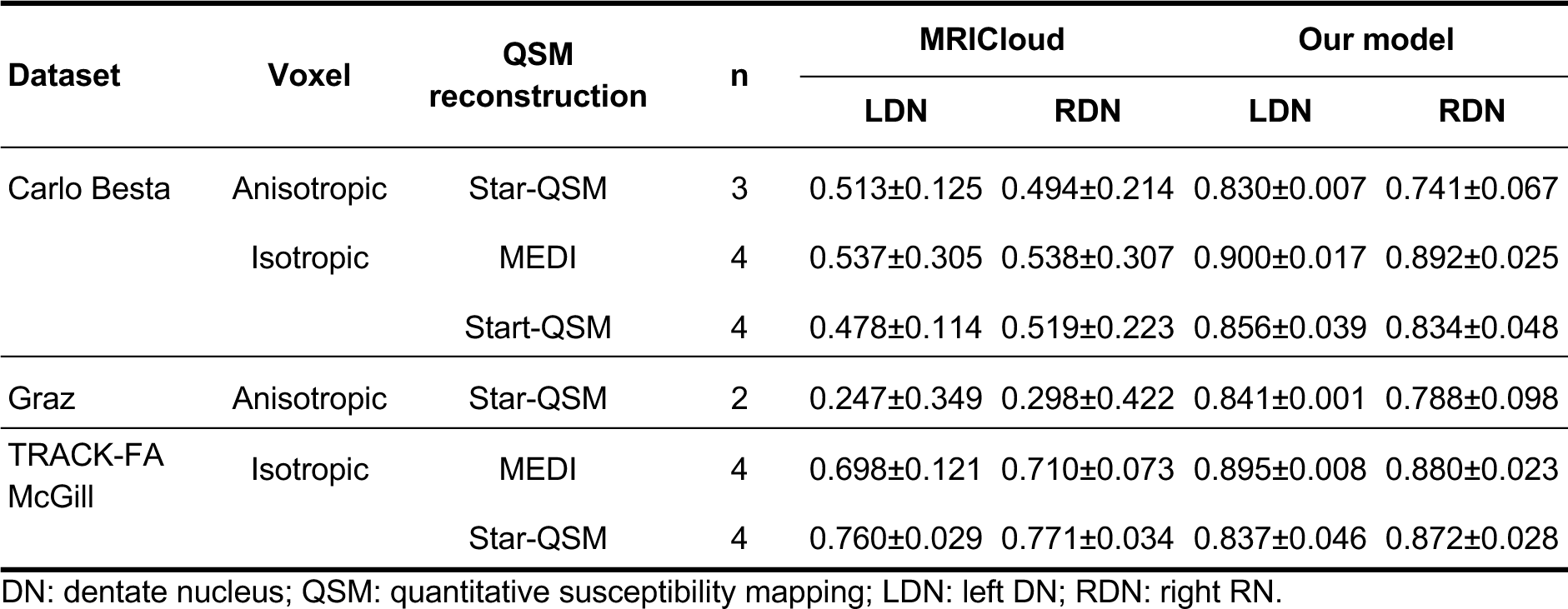
External validation Dice scores (mean±SD) for both DN, comparing results from MRICloud and our proposed DL model.

| Dataset | Voxel | QSM reconstruction | n | MRICloud |  | Our model |  |
| --- | --- | --- | --- | --- | --- | --- | --- |
|  |  |  |  | LDN | RDN | LDN | RDN |
| Carlo Besta | Anisotropic | Star-QSM | 3 | 0.513±0.125 | 0.494±0.214 | 0.830±0.007 | 0.741±0.067 |
| | Isotropic | MEDI | 4 | 0.537±0.305 | 0.538±0.307 | 0.900±0.017 | 0.892±0.025 |
| | | Star-QSM | 4 | 0.478±0.114 | 0.519±0.223 | 0.856±0.039 | 0.834±0.048 |
| Graz | Anisotropic | Star-QSM | 2 | 0.247±0.349 | 0.298±0.422 | 0.841±0.001 | 0.788±0.098 |
| TRACK-FA McGill | Isotropic | MEDI | 4 | 0.698±0.121 | 0.710±0.073 | 0.895±0.008 | 0.880±0.023 |
| | | Star-QSM | 4 | 0.760±0.029 | 0.771±0.034 | 0.837±0.046 | 0.872±0.028 |

DN: dentate nucleus; QSM: quantitative susceptibility mapping; LDN: left DN; RDN: right RN.

**Table 8.** Quantitative validation (mean±SD) of QSM reconstruction methods on external data.

| QSM reconstruction | Voxel | MRICloud |  | Our model |  |
| --- | --- | --- | --- | --- | --- |
|  |  | LDN | RDN | LDN | RDN |
| MEDI | Isotropic | 0.618±0.231 | 0.624±0.226 | 0.898±0.012 | 0.886±0.023 |
| Star-QSM | Anisotropic | 0.407±0.244 | 0.415±0.281 | 0.835±0.008 | 0.760±0.073 |
| | Isotropic | 0.619±0.169 | 0.645±0.200 | 0.847±0.041 | 0.853±0.042 |

DN: dentate nucleus; QSM: quantitative susceptibility mapping; LDN: left DN; RDN: right RN.

### 4.6. Biological Findings

#### 4.6.1. DN Volume versus Mean Magnetic Susceptibility

Significant positive correlations between DN volume and magnetic susceptibility were observed in the healthy control group (r>0.3, Figure 8, Figure S2). These findings were consistently observed in manual tracings (Figure S2) and model-predicted segmentations (Figure 8), with correction for age and head size (total intracranial volume). Moreover, the correlation is stronger in children (<18 years) than in adults. This effect replicates previous reports^19,20^.

**Figure 8.**
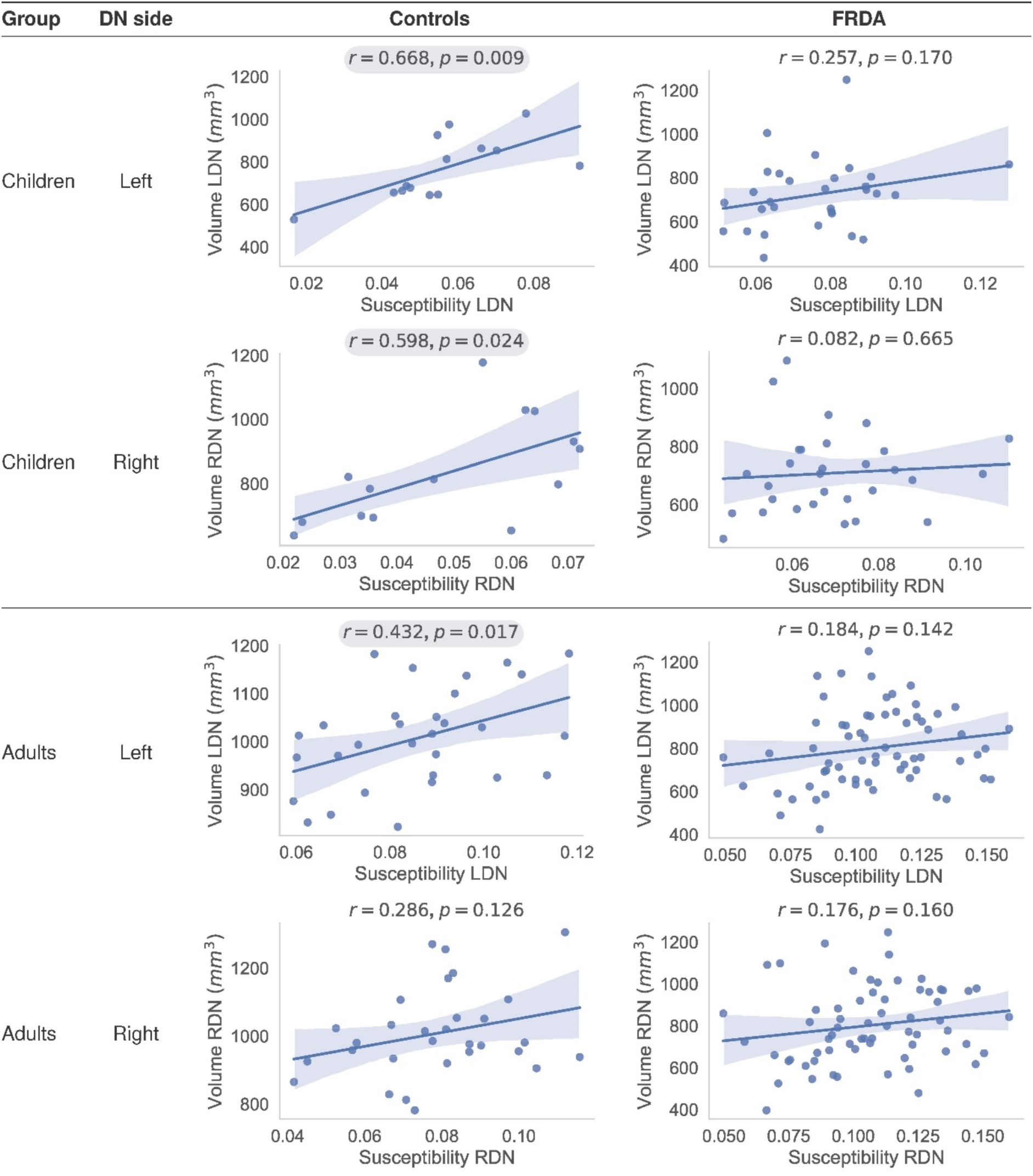
Model prediction results. Pearson’s correlation coefficients and p-values for each group of individuals and DN side. Correlation coefficients with p<0.05 are emphasized with a gray background. Children: subjects under 18 years of age. The volume estimations were corrected for age and head size (eTIV). FRDA: Friedreich’s ataxia; DN: dentate nucleus; LDN: left DN; RDN: right DN.

This correlation suggests that the susceptibility/intensity level may impact border detection due to partial volume effects, possibly introducing a bias in volume estimation. However, the volume versus susceptibility correlation persists, although it is partially attenuated, even after z-score intensity normalization (in a subset of n=20 images, before normalization: LDN (r=0.288), RDN (r=0.197); after normalization: LDN (r=0.087), RDN (r=0.347)).

#### 4.6.2. Statistical Correction for Susceptibility-Driven Volume Bias

The impact of susceptibility variability on volume estimation may be estimated, and thus corrected for, using susceptibility and site (if applicable) as independent variables in multiple linear regression. A correction factor can be estimated from healthy control samples, and subsequently applied to patient data, to avoid confounds between true disease effects and quantification artifacts.

We have estimated a representation correction factor (𝐶𝐹 = 3636.84) in a large cohort of demographically-diverse healthy individuals (age range 11-64 years, male to female ratio of 0.93:1) and several imaging protocols (including different imaging resolutions), which can be used to adjust data in future research studies using the following equation:

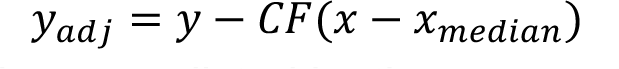

where 𝑦 is the individual DN volume predicted by the segmentation model, 𝑥 is the individual median DN susceptibility in ppm, *x_median_* is the population estimate of median DN susceptibility across a dataset, and 𝐶𝐹 is the regression/correction factor. Note that this formula must be applied uniformly across both disease and control groups in a study to ensure accurate comparative analyses.

## 5. Discussion

Here we propose and develop an automated DN segmentation solution consisting of a two-step deep learning pipeline based on convolutional neural networks. Starting from a comprehensive dataset of QSM images sourced from several MRI centers and high-quality DN manual tracings by experienced annotators, we trained and tested multiple potential model architectures. All tested models delivered statistically superior results compared to MRICloud, the current state-of-the-art automated technique. Our final trained pipeline using the nnU-Net framework performed strongly during external validation, and we have introduced guidance regarding appropriately controlling for the influence of magnetic susceptibility on volume measurements. This work provides a robust, new, open-source tool (https://github.com/art2mri/QSM-Cereb) to the neuroscience community.

In the field of artificial intelligence, validation with external datasets is crucial to assess the generalization capacity of the proposed models^61^, although rarely reported^62^. This step is important to challenge the model across different acquisition protocols, populations, MRI sites, and scanner vendors, providing a valuable way to perform a real-world scenario analysis by evaluating model robustness and performance on unseen data^63^. Our external validation provided strong support for the accuracy and robustness of the model, indicated a low risk of overfitting, and exceeded the accuracy of currently available approaches across a range of experimental conditions (different imaging acquisitions and QSM reconstruction protocols). When focusing on the reconstruction pipeline metrics, after aggregating the results, consistent Dice performance metrics were observed regardless of which QSM pipeline and reconstruction method was used (Table 8). A greater score for MEDI reconstructed images may derive from the fact that experienced raters were exposed to a majority of QSM images processed using this pipeline. We therefore provide a generalizable, fast and open-source solution for accurate and automated DN segmentation.

A combination of several factors contributes to the performance of the proposed model. First, we have gathered an extensive dataset that effectively captures the diversity found in real-world data, enabling good generalizability of the trained model. Additionally, we directly compared multiple DL frameworks to select the nnU-Net^34^ as the optimal architecture for our medical imaging segmentation task among the architectures we tested. The nnU-Net incorporates a well-known architecture and a thorough set of data augmentation techniques. Moreover, our findings are aligned with analogous studies, such as the TotalSegmentator^64^, wherein the nnU-Net stands out as a recognized solution for intricate tasks in the medical imaging domain. Also, our proposed pipeline is composed of two-step models, comprising localization followed by segmentation. In this way, we drive the focus of the second model into a restricted region of the brain, mitigating false positives. Certain voxels in the basal ganglia were predicted as DN in some outputs when the entire image was provided as input, potentially due to the presence of iron in these structures and slight shape similarity. The entire pipeline runs in less than 60 seconds using only the CPU and at most 15 seconds in GPU-based hardware (Intel Core i7 and Nvidia RTX 4070 12 GB).

This work provides a reliable automated solution to a task that has often been undertaken, to-date, by manual delineation (i.e., human hand-drawn segmentation). As such, research outcomes have traditionally relied on the availability of human expertise and are prone to errors resulting from task fatigue and inherent human variability. These considerations have limited the efficiency and accuracy of research outcomes, such as volumetric studies of DN volume in populations with neurological disorders that impact the cerebellum. The availability of a fast, accurate, and scalable automated segmentation tool opens new avenues of research into the DN structure in health and disease, including longitudinal natural history studies and clinical trials in disorders with known DN involvement, such as FRDA and SCA3^65^.

In terms of biological findings, we replicated previously reported positive significant correlations between dentate nucleus volume and the magnetic susceptibility measures, in the healthy population^19,20^. These effects were present both in the manual tracings and the prediction masks obtained using the trained models and could be explained by partial volume effects along the edges of the structure, whereby voxels that include a mix of tissue both inside and outside the DN would be more likely to be identified as being inside the DN in individuals with higher mean susceptibility intensity. The result persisted (albeit with a lesser magnitude) even after intensity normalization. Moreover, work by Li and colleagues^19^ also found that this positive association (with manual tracing) was present in the DN, but not in other midbrain or basal ganglia nuclei. If intensity-induced variability were indeed driving partial volume effects, this effect would be expected to be more severe in small structures such as the DN and the red nucleus^19^.

Regardless of the mechanism (biological or methodological) underlying this dependency between susceptibility and volume, it could be seen as an artifact that may be accounted for in DN volume assessments. This is further supported by the observation that, because susceptibility increases naturally with age, failure to correct for this dependency results in an apparent positive relationship between DN volume and age in the adult population (i.e., apparent DN growth over time)^19,20^, which is biologically implausible. Care must also be taken when assessing populations with brain pathology, as the relationship between susceptibility and volume in neurological conditions may be a mix of true disease effects and the artifact described above. Neurological diseases, such as FRDA, SCA1, and multiple system atrophy are characterized by increased DN susceptibility^8,66^, which may lead to underestimation of DN volume loss. We, therefore, recommend statistically controlling for susceptibility levels when inferring volume effects to account for this artifact. In studies of normative populations, this can be achieved through simple regression (e.g., including a predictor of non-interest coding for susceptibility in regression models); for studies in pathological conditions, a correction factor estimated from normative data can be applied.

We have reported a correction factor of 3636.84 for adjusting the total volume of DN, which represents the average of LDN and RDN volumes. However, it is important to note that slightly different correction factors were observed for LDN (3553.7188) and RDN (3422.2106). Consequently, future studies may select the appropriate correction factor based on their specific data and objective. Additionally, adjustments for other confounding variables, such as the impact of aging on DN volumes, susceptibility, or other measures, can be made either in sequential steps or by integrating all the confounding variables into a multiple regression model.

A limitation of this study lies in the fact that only three QSM reconstruction pipelines were used for generating the training images. Several QSM pipelines are available, including the possibility of combining a wide range of phase unwrapping, background field removal, and susceptibility mapping reconstruction methods and parameters. However, collecting such a diverse dataset would impose another layer of complexity on the study. As the QSM field continues to mature, it is likely that a narrower range of acquisition and reconstruction protocols will become the norm, reducing variability in this dimension. Notably, recent consensus papers have provided recommendations focused on clinical research in this area^67,68^, which might be usefully explored in future investigations. Future work should also explore histological validation of the imaging segmentations to provide further improved ground truth. Similarly, higher resolution imaging protocols combined with increasingly accurate QSM reconstruction approaches offer the potential to isolate the gray matter ribbon of the DN from the central white matter core, which will enable tissue-type specific inferences.

In conclusion, our work provides state-of-the-art performance in automated DN segmentation from *in vivo* MRI based on extensive training and evaluation of a diverse dataset and methods. This outcome provides an important tool for characterizing cerebellar neuroanatomy in health and disease and biomarker discovery relevant for tracking disease progression and treatment efficacy in cerebellar disorders.

## 6. Data and Code Availability Statement

The patient MRI data are not publicly available due to privacy regulations. Access can be provided upon reasonable request to scientists in accordance with our Data Use and Access Policy. TRACK-FA and Enigma data might be provided upon request directed to Helena Bujalka or Ian Harding.

The source code of QSM deep-learning segmentation pipeline will be made publicly available on GitHub (https://github.com/art2mri/QSM-Cereb) upon acceptance.

## Data Availability

The patient MRI data is not publicly available due to privacy regulations. Access can be provided upon reasonable request to scientists in accordance with our Data Use and Access Policy.

## Acknowledgments

The authors gratefully thank all of the individuals for their participation in the study and the Friedreich’s Ataxia Research Alliance (FARA, USA) for their help with FRDA participant recruitment.

The authors thank and acknowledge the FARA, Takeda Pharmaceuticals Company, Novartis Gene Therapies, IXICO plc and PTC Therapeutics for support in funding and for their participation in the TRACK-FA Neuroimaging Consortium, including providing advice and feedback on study design, implementation and analysis of outcomes.

Study data were collected and managed using REDCap electronic data capture tools hosted and managed by Helix (Monash University, Australia).

Study data were collected, stored, and managed using XNAT, a software framework for managing neuroimaging laboratory data hosted by Monash (MXNAT) and Monash Biomedical Imaging (MBIXNAT) (Monash University, Australia).

The authors thank the research coordinators across all TRACK-FA sites for their assistance in participant recruitment and testing.

The following TRACK-FA collaborators opted to have their name acknowledged: Helena Bujalka, School of Psychological Sciences, The Turner Institute for Brain and Mental Health, Monash University, Clayton, Victoria, Australia; Manuela Corti, Powell Gene Therapy Centre, University of Florida, Gainesville, Florida, United States of America; Jennifer Farmer, Friedreich’s Ataxia Research Alliance (FARA), Downingtown, Pennsylvania, United States of America; Myriam Rai, Friedreich’s Ataxia Research Alliance (FARA), Downingtown, Pennsylvania, United States of America.

## Author contributions

Cohort principal investigator (PI): TJRR, IHH, MCFJ, SC, MBD, ID, NGK, PGH, CL, MP, KR. Imaging data analysis: DHS, SS, TJRR, IHH, IMA, JMJ, CCL, SM, AS. Imaging data collection: TJRR, MCFJ, SC, LAC, ID, MG, PGH, JMJ, CLK, THM, AN, MP, KR, TPR, SR, DAR. Clinical data collection: TJRR, MCFJ, LAC, ID, CLK, ARMM, MP, KR, SHS. Intellectual contribution to study design & conduct at your site: DHS, TJRR, IHH, SC, ID, WG, NGK, PGH, GMJ, ARMM, MP, TPR, DAR, JBS, SHS. Methods development: DHS, TJRR, IHH, NGK, SG. Statistical analysis: DHS, SS, TJRR, EFL. Manuscript review: DHS, SS, TJRR, IHH, MCFJ, SC, LAC, AD, ID, WG, NGK, SG, MG, PGH, CLK, CL, JL, EFL, THM, ARMM, AN, KR, TPR, SR, DAR, JBS, SHS, DT.

## Funding

This work was supported by the Friedreich’s Ataxia Research Alliance (FARA General Research Grant) and grants from the Australian National Health and Medical Research Council (NHMRC Ideas Grant 1184403). The funding agencies did not influence the study design, data collection, or manuscript drafting.

## Competing interests

TJRR is employed by Biogen, receives a salary, and is a grant recipient from Friedreich’s Ataxia Research Alliance (FARA). DHS is an Itaú Unibanco SA employee and a grant recipient from Friedreich’s Ataxia Research Alliance (FARA). SC received fees from Amicus for the advisory board. LAC is funded by Friedreich’s Ataxia Research Alliance (FARA) and is a consultant for Biogen Pharmaceuticals. ID is funded by Friedreich’s Ataxia Research Alliance (FARA). NGK is funded by Friedreich’s Ataxia Research Alliance (FARA) and CHDI Foundation Inc (New York, USA). PGH is a grant recipient from Friedreich’s Ataxia Research Alliance (FARA) and the National Institute of Health (NIH) P41EB027061, P30NS076408, S10OD017974. JMJ is funded by Friedreich’s Ataxia Research Alliance (FARA) and National Institute of Health (NIH). CLK reports grant FWF P35887. CL received research grants from Minoryx Therapeutics, research support from Biogen Inc and is funded by Friedreich’s Ataxia Research Alliance (FARA) and NIH P41 EB027061. JL is funded by Friedreich’s Ataxia Research Alliance (FARA). TM reports funding from National Institute of Health (NIH) U01 NS104326. SM is funded by the Italian MUR for the project “SEE LIFE - StrEngthEning the Italian InFrastructure of Euro-bioimaging.” AN is funded by the Italian Ministry of Health (RRC). KR is funded by Friedreich’s Ataxia Research Alliance (FARA). SHS is industry support research for Reata, Biogen, Biohaven, Avidity Biosciences, Fulcrum therapeutics, Vertex, Arthex, PTC, Reneo, Larimar, has a consulting work for Reata, Biogen, Fulcrum, participated in speaking engagements for Biogen, MDA, Bionews, Medscape, and is funded by NIH, FDA, Muscular Dystrophy Association, Wyck Foundation, Friedreich’s Ataxia Research Alliance, and National Ataxia Foundation. DT reports grants DFG, DE 2516/1-1 and TI 239/17-1. MCFJ reports funding from FAPESP (São Paulo Research Foundation). IHH reports Friedreich’s Ataxia Research Alliance general research grant and NHMRC Ideas and Investigator Grants (2026191, 1184403). All other authors declare no financial or non-financial competing interests.

## Notes

### Author Declarations

The ethics committee or institutional review board (IRB) respective to each project data source or site approved the use or ethics waiver for this retrospective study, and all participants provided written informed consent prior to original data collection. The TRACK-FA steering committee approved the data use, and IRB reference numbers were previously published (Monash Health Human Research Ethics Committee: RES-20-0000-139A; Children's Hospital of Philadelphia: IRB 20-017611; University of Minnesota: IRB STUDY00009047; University of Florida: IRB202000399; RWTH Aachen University: EK195/20; University of Campinas (CAAE NO): 83241318.3.1001.5404; McGill IRB Approved Project Number: 2022-8676). Ethics approval was obtained independently for the remaining studies, respectively: Ethical Committee "Carlo Romano" of the University of Naples "Federico II" (Naples A: 209/13, Naples B: 47/15), Monash University Human Research Ethics Committee (IMAGE-FRDA: 13201B, INFLAM-FRDA: 7810), and University of Minnesota IRB (1210M22281). The institutional ethics committee respective to each project approved their study (Fondazione IRCCS Istituto Neurologico "Carlo Besta": 42/2017 07/06/2017; Medical University of Graz local ethics-committee: 31-432ex18/191264-2019).

### Summary of Updates

The section on Swin UNETR was updated to correct the architecture definition and fundamentals.

## References

1 Dum, R. P. & Strick, P. L. An unfolded map of the cerebellar dentate nucleus and its projections to the cerebral cortex. J Neurophysiol 89, 634–639 (2003). 10.1152/jn.00626.2002

2 Henschke, J. U. & Pakan, J. M. Disynaptic cerebrocerebellar pathways originating from multiple functionally distinct cortical areas. Elife 9 (2020). 10.7554/eLife.59148

3 Stoodley, C. J. & Schmahmann, J. D. Evidence for topographic organization in the cerebellum of motor control versus cognitive and affective processing. Cortex 46, 831–844 (2010). 10.1016/j.cortex.2009.11.008

4 Anteraper, S. A. et al. Intrinsic Functional Connectivity of Dentate Nuclei in Autism Spectrum Disorder. Brain Connect 9, 692–702 (2019). 10.1089/brain.2019.0692

5 Olivito, G. et al. Cerebellar dentate nucleus functional connectivity with cerebral cortex in Alzheimer’s disease and memory: a seed-based approach. Neurobiol Aging 89, 32–40 (2020). 10.1016/j.neurobiolaging.2019.10.026

6 Xie, Y. J. et al. Functional connectivity of cerebellar dentate nucleus and cognitive impairments in patients with drug-naive and first-episode schizophrenia. Psychiatry Res 300, 113937 (2021). 10.1016/j.psychres.2021.113937

7 Koeppen, A. H. The pathogenesis of spinocerebellar ataxia. Cerebellum 4, 62–73 (2005). 10.1080/14734220510007950

8 Deistung, A. et al. Quantitative susceptibility mapping reveals alterations of dentate nuclei in common types of degenerative cerebellar ataxias. Brain Commun 4, fcab306 (2022). 10.1093/braincomms/fcab306

9 Jaschke, D. et al. Age-related differences of cerebellar cortex and nuclei: MRI findings in healthy controls and its application to spinocerebellar ataxia (SCA6) patients. Neuroimage 270, 119950 (2023). 10.1016/j.neuroimage.2023.119950

10 Stefanescu, M. R. et al. Structural and functional MRI abnormalities of cerebellar cortex and nuclei in SCA3, SCA6 and Friedreich’s ataxia. Brain 138, 1182–1197 (2015). 10.1093/brain/awv064

11 Ward, P. G. D. et al. Longitudinal evaluation of iron concentration and atrophy in the dentate nuclei in friedreich ataxia. Mov Disord 34, 335–343 (2019). 10.1002/mds.27606

12 Diedrichsen, J. et al. Imaging the deep cerebellar nuclei: a probabilistic atlas and normalization procedure. Neuroimage 54, 1786–1794 (2011). 10.1016/j.neuroimage.2010.10.035

13 Haacke, E. M., Xu, Y., Cheng, Y. C. & Reichenbach, J. R. Susceptibility weighted imaging (SWI). Magn Reson Med 52, 612–618 (2004). 10.1002/mrm.20198

14 Ramos, P. et al. Iron levels in the human brain: a post-mortem study of anatomical region differences and age-related changes. J Trace Elem Med Biol 28, 13–17 (2014). 10.1016/j.jtemb.2013.08.001

15 Halefoglu, A. M. & Yousem, D. M. Susceptibility weighted imaging: Clinical applications and future directions. World J Radiol 10, 30–45 (2018). 10.4329/wjr.v10.i4.30

16 Liu, C., Li, W., Tong, K. A., Yeom, K. W. & Kuzminski, S. Susceptibility-weighted imaging and quantitative susceptibility mapping in the brain. J Magn Reson Imaging 42, 23–41 (2015). 10.1002/jmri.24768

17 Langkammer, C. et al. Quantitative susceptibility mapping (QSM) as a means to measure brain iron? A post mortem validation study. Neuroimage 62, 1593–1599 (2012). 10.1016/j.neuroimage.2012.05.049

18 Bonilha da Silva, C., et al. Dentate nuclei T2 relaxometry is a reliable neuroimaging marker in Friedreich’s ataxia. Eur J Neurol 21, 1131–1136 (2014). 10.1111/ene.12448

19 Li, G. et al. Age-dependent changes in brain iron deposition and volume in deep gray matter nuclei using quantitative susceptibility mapping. Neuroimage 269, 119923 (2023). 10.1016/j.neuroimage.2023.119923

20 Zhang, Y. et al. Visualizing the deep cerebellar nuclei using quantitative susceptibility mapping: An application in healthy controls, Parkinson’s disease patients and essential tremor patients. Hum Brain Mapp 44, 1810–1824 (2023). 10.1002/hbm.26178

21 Li, X. et al. Multi-atlas tool for automated segmentation of brain gray matter nuclei and quantification of their magnetic susceptibility. Neuroimage 191, 337–349 (2019). 10.1016/j.neuroimage.2019.02.016

22 Mori, S. et al. MRICloud: delivering high-throughput MRI neuroinformatics as cloud-based software as a service. Computing in Science & Engineering 18, 21–35 (2016).

23 Georgiou-Karistianis, N. et al. A natural history study to track brain and spinal cord changes in individuals with Friedreich’s ataxia: TRACK-FA study protocol. PLoS One 17, e0269649 (2022). 10.1371/journal.pone.0269649

24 Khan, W. et al. Neuroinflammation in the Cerebellum and Brainstem in Friedreich Ataxia: An [18F]-FEMPA PET Study. Mov Disord 37, 218–224 (2022). 10.1002/mds.28825

25 Chen, L. et al. Quantitative Susceptibility Mapping of Brain Iron and beta-Amyloid in MRI and PET Relating to Cognitive Performance in Cognitively Normal Older Adults. Radiology 298, 353–362 (2021). 10.1148/radiol.2020201603

26 Fang, J., Bao, L., Li, X., van Zijl, P. C. M. & Chen, Z. Background field removal using a region adaptive kernel for quantitative susceptibility mapping of human brain. J Magn Reson 281, 130–140 (2017). 10.1016/j.jmr.2017.05.004

27 Liu, T. et al. Morphology enabled dipole inversion (MEDI) from a single-angle acquisition: comparison with COSMOS in human brain imaging. Magn Reson Med 66, 777–783 (2011). 10.1002/mrm.22816

28 Liu, J. et al. Morphology enabled dipole inversion for quantitative susceptibility mapping using structural consistency between the magnitude image and the susceptibility map. Neuroimage 59, 2560–2568 (2012). 10.1016/j.neuroimage.2011.08.082

29 Li, W., Wu, B. & Liu, C. Quantitative susceptibility mapping of human brain reflects spatial variation in tissue composition. Neuroimage 55, 1645–1656 (2011). 10.1016/j.neuroimage.2010.11.088

30 Yushkevich, P. A. et al. User-guided 3D active contour segmentation of anatomical structures: significantly improved efficiency and reliability. Neuroimage 31, 1116–1128 (2006). 10.1016/j.neuroimage.2006.01.015

31 FSLeyes v. 1.11.0 (Zenodo, 2024).

32 Ronneberger, O., Fischer, P. & Brox, T. in Medical image computing and computer-assisted intervention–MICCAI 2015: 18th international conference, Munich, Germany, October 5-9, 2015, proceedings, part III 18. 234–241 (Springer).

33 Çiçek, Ö., Abdulkadir, A., Lienkamp, S. S., Brox, T. & Ronneberger, O. in Medical Image Computing and Computer-Assisted Intervention–MICCAI 2016: 19th International Conference, Athens, Greece, October 17-21, 2016, Proceedings, Part II 19. 424–432 (Springer).

34 Isensee, F., Jaeger, P. F., Kohl, S. A. A., Petersen, J. & Maier-Hein, K. H. nnU-Net: a self-configuring method for deep learning-based biomedical image segmentation. Nat Methods 18, 203–211 (2021). 10.1038/s41592-020-01008-z

35 Lee, C.-Y., Xie, S., Gallagher, P., Zhang, Z. & Tu, Z. in Artificial intelligence and statistics. 562-570 (Pmlr).

36 Vaswani, A. et al. Attention is all you need. Advances in neural information processing systems 30 (2017).

37 Chen, J., et al. Transunet: Transformers make strong encoders for medical image segmentation. *arXiv preprint arXiv:2102.04306* (2021).

38 Hatamizadeh, A. et al. in Proceedings of the IEEE/CVF winter conference on applications of computer vision. 574-584.

39 Hatamizadeh, A. et al. in International MICCAI Brainlesion Workshop. 272-284 (Springer).

40 Liu, Z. et al. in Proceedings of the IEEE/CVF international conference on computer vision. 10012–10022.

41 Dosovitskiy, A., et al. An image is worth 16×16 words: Transformers for image recognition at scale. *arXiv preprint arXiv:2010.11929* (2020).

42 Cardoso, M. J., et al. Monai: An open-source framework for deep learning in healthcare. *arXiv preprint arXiv:2211.02701* (2022).

43 Han, S., Carass, A., He, Y. & Prince, J. L. Automatic cerebellum anatomical parcellation using U-Net with locally constrained optimization. Neuroimage 218, 116819 (2020). 10.1016/j.neuroimage.2020.116819

44 Avants, B. B. et al. A reproducible evaluation of ANTs similarity metric performance in brain image registration. Neuroimage 54, 2033–2044 (2011). 10.1016/j.neuroimage.2010.09.025

45 Dice, L. R. Measures of the Amount of Ecologic Association between Species. Ecology 26, 297–302 (1945). Doi 10.2307/1932409

46 Sorenson, T. A method of establishing groups of equal amplitude in plant sociology based on similarity of species content, and its application to analysis of vegetation on Danish commons. Kong Dan Vidensk Selsk Biol Skr 5, 1–5 (1948).

47 Bertels, J. et al. Optimizing the Dice Score and Jaccard Index for Medical Image Segmentation: Theory and Practice. Lect Notes Comput Sc 11765, 92–100 (2019). 10.1007/978-3-030-32245-8_11

48 Huttenlocher, D. P., Klanderman, G. A. & Rucklidge, W. J. Comparing Images Using the Hausdorff Distance. Ieee T Pattern Anal 15, 850–863 (1993). Doi 10.1109/34.232073

49 Taha, A. A. & Hanbury, A. Metrics for evaluating 3D medical image segmentation: analysis, selection, and tool. BMC Med Imaging 15, 29 (2015). 10.1186/s12880-015-0068-x

50 Jaccard, P. The distribution of the flora in the alpine zone. 1. New phytologist 11, 37–50 (1912).

51 Koo, T. K. & Li, M. Y. A Guideline of Selecting and Reporting Intraclass Correlation Coefficients for Reliability Research. J Chiropr Med 15, 155–163 (2016). 10.1016/j.jcm.2016.02.012

52 Jenkinson, M., Beckmann, C. F., Behrens, T. E., Woolrich, M. W. & Smith, S. M. Fsl. Neuroimage 62, 782–790 (2012). 10.1016/j.neuroimage.2011.09.015

53 van Bergen, J. M. et al. Quantitative Susceptibility Mapping Suggests Altered Brain Iron in Premanifest Huntington Disease. AJNR Am J Neuroradiol 37, 789–796 (2016). 10.3174/ajnr.A4617

54 Chan, K. S. & Marques, J. P. SEPIA-Susceptibility mapping pipeline tool for phase images. Neuroimage 227, 117611 (2021). 10.1016/j.neuroimage.2020.117611

55 Abdul-Rahman, H. S. et al. Fast and robust three-dimensional best path phase unwrapping algorithm. Appl Opt 46, 6623–6635 (2007). 10.1364/ao.46.006623

56 Zhou, D., Liu, T., Spincemaille, P. & Wang, Y. Background field removal by solving the Laplacian boundary value problem. NMR Biomed 27, 312–319 (2014). 10.1002/nbm.3064

57 Wei, H. et al. Streaking artifact reduction for quantitative susceptibility mapping of sources with large dynamic range. NMR Biomed 28, 1294–1303 (2015). 10.1002/nbm.3383

58 Ulyanov, D., Vedaldi, A. & Lempitsky, V. Instance normalization: The missing ingredient for fast stylization. *arXiv preprint arXiv:1607.08022* (2016).

59 He, K., Zhang, X., Ren, S. & Sun, J. in Proceedings of the IEEE international conference on computer vision. 1026–1034.

60 Loshchilov, I. & Hutter, F. Decoupled weight decay regularization. *arXiv preprint arXiv:1711.05101* (2017).

61 Ramspek, C. L., Jager, K. J., Dekker, F. W., Zoccali, C. & van Diepen, M. External validation of prognostic models: what, why, how, when and where? Clin Kidney J 14, 49–58 (2021). 10.1093/ckj/sfaa188

62 Collins, G. S. & Moons, K. G. M. Reporting of artificial intelligence prediction models. Lancet 393, 1577–1579 (2019). 10.1016/S0140-6736(19)30037-6

63 de Hond, A. A. H. et al. Guidelines and quality criteria for artificial intelligence-based prediction models in healthcare: a scoping review. NPJ Digit Med 5, 2 (2022). 10.1038/s41746-021-00549-7

64 Wasserthal, J. et al. TotalSegmentator: Robust Segmentation of 104 Anatomic Structures in CT Images. Radiol Artif Intell 5, e230024 (2023). 10.1148/ryai.230024

65 Koeppen, A. H. The Neuropathology of Spinocerebellar Ataxia Type 3/Machado-Joseph Disease. Adv Exp Med Biol 1049, 233–241 (2018). 10.1007/978-3-319-71779-1_11

66 Harding, I. H. et al. Tissue atrophy and elevated iron concentration in the extrapyramidal motor system in Friedreich ataxia: the IMAGE-FRDA study. J Neurol Neurosurg Psychiatry 87, 1261–1263 (2016). 10.1136/jnnp-2015-312665

67 Committee, Q. S. M. C. O. et al. Recommended implementation of quantitative susceptibility mapping for clinical research in the brain: A consensus of the ISMRM electro-magnetic tissue properties study group. Magn Reson Med 91, 1834–1862 (2024). 10.1002/mrm.30006

68 Oz, G. et al. MR Imaging in Ataxias: Consensus Recommendations by the Ataxia Global Initiative Working Group on MRI Biomarkers. Cerebellum (2023). 10.1007/s12311-023-01572-y

