## Supplementary Material for "Automated Dentate Nucleus Segmentation from QSM Images Using Deep Learning"

* *Contributed equally*
† *Senior principal investigator*

#### Deep learning model training specifications

The models explored in the architecture experimentation task included U-Net[1] with deep supervision (DS)[2], Swin UNETR[3], and the nnU-Net framework[4]. U-Net with DS started with feature channels at 32, doubling up to a value of 512. Each level combined two 3x3x3 convolutions, followed by instance normalization[5] and Leaky Rectified Linear Unit (Leaky ReLU)[6] activation with a slope of 0.01. A Stochastic Gradient Descent (SGD) optimizer with a learning rate of $3x10^{-2}$ and a Nesterov momentum[7,8] of 0.99 was used, combined with a Dice loss function for training. The learning rate was decayed using a polynomial function scheduler. Training lasted 600 epochs with a batch size of 1. The original Swin UNETR implementation was configured with a feature size of 24. An AdamW[9] optimizer was selected with a learning rate of $1x10^{-4}$, weight decay of $1x10^{-5}$, $\beta 1 = 0.9$, $\beta 2 = 0.999$, and $\epsilon = 1x10^{-8}$. For the loss function, we used the Dice loss. A cosine annealing learning rate scheduler was set. The network was trained for 1000 epochs and a batch size of 1. Finally, the nnU-Net framework was trained using the default settings for the 3D full resolution trainer. For both U-Net with DS and Swin UNETR, a same set of augmentation techniques were selected. To replicate the nnU-Net augmentation pipeline, we incorporated random rotation, shear, scaling, translation, flipping, elastic deformation, and gamma contrast methods.

### External Validation

**Table S1**. External validation datasets acquisition parameters.

| Dataset | Scanner | Sequence | TR (ms) | TE1 (ms) | $\Delta$TE (ms) | # of echoes | FoV (mm) | Image matrix (voxels) | Voxel size (mm) | Acquisition time |
|---|---|---|---|---|---|---|---|---|---|---|
| TRACK-FA McGill | 3T Siemens Prisma | GRE | 27 | 3.7 | 6 | 4 | 220 x 220 x 176 | 208 x 256 x 176 | 0.86 iso | 7' 22" |
| Carlo Besta | 3T Philips Achieva | GRE | 40 | 4.5 | 5 | 7 | 240 x 180<br>140 axial slices | 480 x 480 x 140 | 0.50 x 0.50 x 1.0 | 4' 23" |
| | 3T Philips Achieva | GRE | 40 | 5.4 | 5.2 | 7 | 224 x 224<br>140 axial slices | 224 x 224 x 140 | 1.0 iso | 8' 12" |
| Graz | 3T Siemens Magnetom Trio | GRE | 68 | 4.92 | 4.92 | 12 | 188 x 230 x 128 | 208 x 256 x 64 | 0.90 x 0.90 x 2.0 | 4' 51" |

GRE: gradient recalled echo; iso: isotropic; TR: repetition time; TE: echo time; TE1: first echo time; FoV: field of view; iso: isotropic.

**Table S2**. External validation datasets demographics.

| | McGill | | Carlo Besta | Graz |
|---|---|---|---|---|
| | **Controls** | **FRDA** | **Controls** | **MS** |
| **Subjects** | 3 | 1 | 4 | 2 |
| **Age** | 29.8±4.4 | 41 | 64.5±10.0 | 43.5±17.7 |
| **Sex (M/F)** | 2/1 | 0/1 | 1/3 | 1/1 |

FRDA: Friedreich's ataxia; MS: multiple sclerosis.

Ethics approval was obtained for the CMRR study: IRB 1210M22281 (University of Minnesota).

**Table S3**. Acquisition protocols for CMRR dataset.

| Dataset | Scanner | Sequence | TR (ms) | TE (ms) | $\Delta$TE (ms) | # of echoes | FoV (mm) | Image matrix (voxels) | Voxel size (mm) | Acquisition time |
|---|---|---|---|---|---|---|---|---|---|---|
| CMRR | 3T Siemens Magnetom Prisma Fit | GRE | 86 | 20.48/30/45 | - | 3 | 205 x 186 x 32 | 232 x 256 x 32 | 0.80 iso | 6' 59" |
| | | | 65 | 7.26 (TE1) | 5.00 | 8 | 205 x 185 x (30-36) | 232 x 256 x (30-36) | 0.80 iso | 4' 40" - 5' 50" |
| | | | 65 | 7.26 (TE1) | 5.00 | 8 | 174 x 192 x 36 | 174 x 192 x 36 | 1.0 iso | 4' 38" |
| | | | 54 | 9.84 (TE1) | 9.84 | 5 | 230 x 230 x 32 | 256 x 256 x 32 | 0.90 iso | 4' 34" |

MRI: magnetic resonance imaging; GRE: gradient recalled echo; iso: isotropic; TR: repetition time; TE: echo time; TE1: first echo time; FoV: field of view; iso: isotropic.

**Table S4**. Subject demographics for CMRR dataset.

| | CMRR | |
|---|---|---|
| | **Controls** | **FRDA** |
| **Subjects** | 3 | 16 |
| **Age** | 25.0±11.1 | 23.6±9.0 |
| **Sex (M/F)** | 2/1 | 6/10 |

FRDA: Friedreich's ataxia.

#### Segmentation Model Architectures Comparison

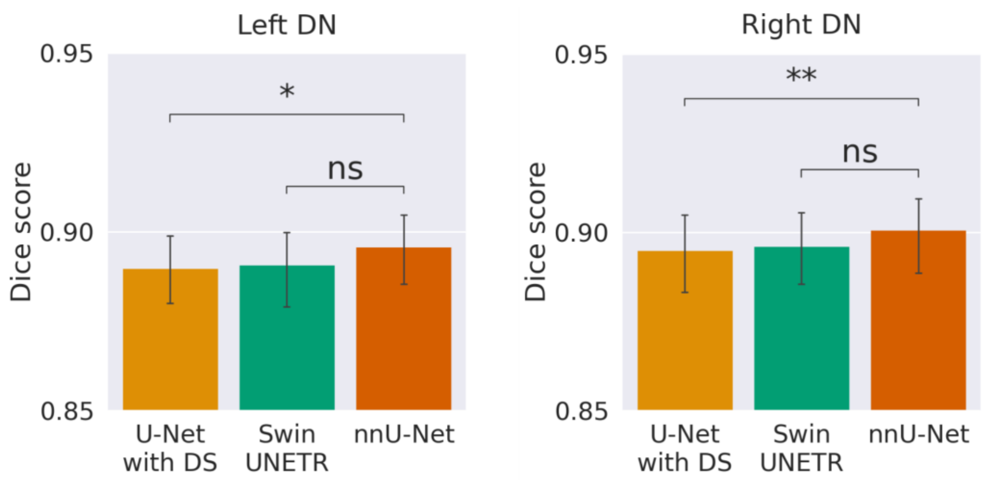


**Figure S1.** Bar plots of trained segmentation models. nnU-Net Dice score is statistically significantly higher than U-Net, and no significance was found when compared to Swin UNETR. ns: non-significant; * $p < 0.05$; ** $p < 0.01$; *** $p < 0.001$; **** $p < 0.0001$.

### DN Volume versus Mean Magnetic Susceptibility

| Group | DN side | Controls | FRDA |
|---|---|---|---|
| Children | Left | 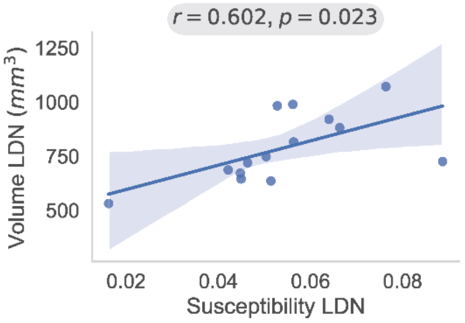 | 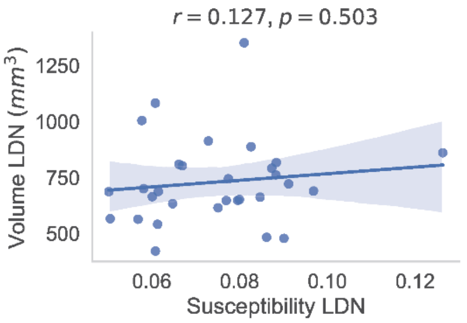 |
| Children | Right | 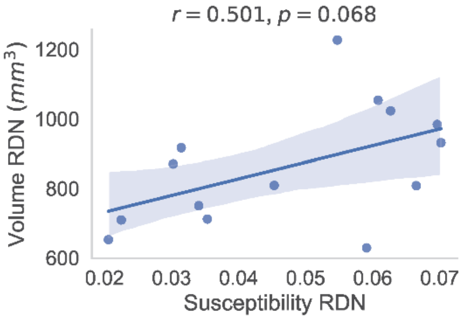 | 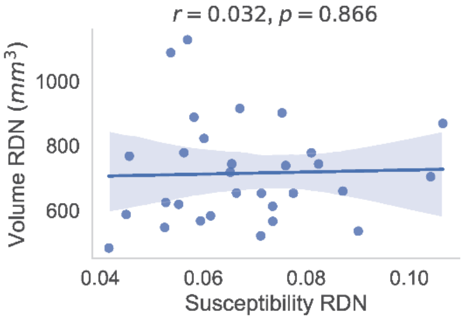 |
| Adults | Left | 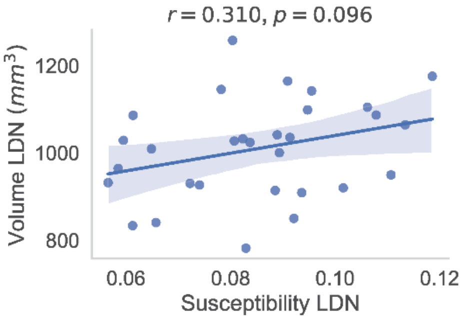 | 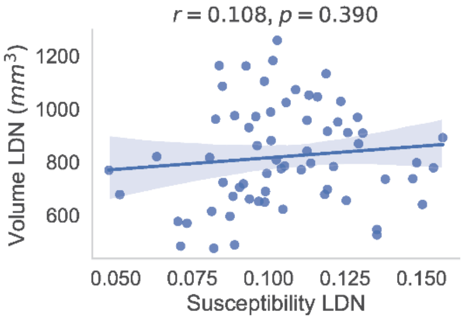 |
| Adults | Right | 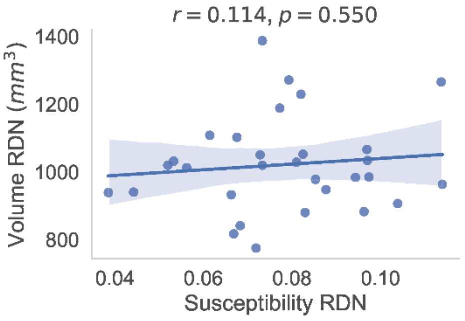 | 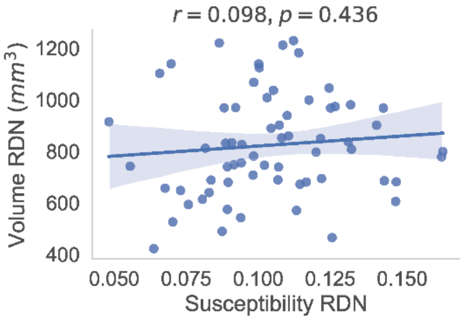 |

**Figure S2.** Manual segmentation results. Pearson’s correlation coefficients and p-values for each group of individuals and DN side. Correlation coefficients with p<0.05 are emphasized with a gray background. Children: subjects under 18 years of age. The volume estimations were corrected for age and head size (eTIV). FRDA: Friedreich’s ataxia; DN: dentate nucleus; LDN: left DN; RDN: right DN; Vol: volume.

### References


1 Çiçek, Ö., Abdulkadir, A., Lienkamp, S. S., Brox, T. & Ronneberger, O. in *Medical Image Computing and Computer-Assisted Intervention–MICCAI 2016: 19th International Conference, Athens, Greece, October 17-21, 2016, Proceedings, Part II 19.* 424-432 (Springer).

2 Lee, C.-Y., Xie, S., Gallagher, P., Zhang, Z. & Tu, Z. in *Artificial intelligence and statistics.* 562-570 (Pmlr).

3 Hatamizadeh, A. *et al.* in *International MICCAI Brainlesion Workshop.* 272-284 (Springer).

4 Isensee, F., Jaeger, P. F., Kohl, S. A. A., Petersen, J. & Maier-Hein, K. H. nnU-Net: a self-configuring method for deep learning-based biomedical image segmentation. *Nat Methods* **18**, 203-211 (2021). https://doi.org/10.1038/s41592-020-01008-z

5 Ulyanov, D., Vedaldi, A. & Lempitsky, V. Instance normalization: The missing ingredient for fast stylization. *arXiv preprint arXiv:1607.08022* (2016).

6 Maas, A. L., Hannun, A. Y. & Ng, A. Y. in *Proc. icml.* 3 (Atlanta, GA).

7 Sutskever, I., Martens, J., Dahl, G. & Hinton, G. in *International conference on machine learning.* 1139-1147 (PMLR).

8 Nesterov, Y. A method of solving a convex programming problem with convergence rate O (1/k** 2). *Doklady Akademii Nauk SSSR* **269**, 543 (1983).

9 Loshchilov, I. & Hutter, F. Decoupled weight decay regularization. *arXiv preprint arXiv:1711.05101* (2017).
